# Quantified Brain Atrophy and Risk of Severe Mass Effect in Acute Ischemic Stroke

**DOI:** 10.64898/2026.02.27.26346805

**Authors:** Yili Du, Aiman Z. Altaf, Nawal J. Ibrahim, Stefanos Chatzidakis, Leigh Ann Mallinger, Allyson L. Reinert, Rebecca A. Stafford, Atul Kumar, Amrit Avula, Mohamad Abdalkader, Huimin Cheng, David M. Greer, Rajat Dhar, Charlene J. Ong

**Affiliations:** Department of Neurology, Boston Medical Center and Boston University Chobanian and Avedisian School of Medicine, Boston, MA, USA; Department of Biostatistics, Boston University School of Public Health, Boston, MA, USA; Department of Neurology, Brigham and Women’s Hospital, Harvard Medical School, Boston, MA, USA; Department of Neurology, Washington University School of Medicine, St. Louis, MO, USA; Department of Radiology, Boston Medical Center and Boston University Chobanian and Avedisian School of Medicine, Boston, MA, USA; Department of Neurosurgery, Boston Medical Center and Boston University Chobanian and Avedisian School of Medicine, Boston, MA, USA

**Author notes:** **Corresponding Author:** Charlene J. Ong, MD, MPHS, Associate Professor, Department of Neurology, Boston University School of Medicine, Boston Medical Center. 85 E Concord St., Suite 1116, Boston, MA 02118.

**Keywords:** Quantified Brain Atrophy, Acute Ischemic Stroke, Cerebral Edema

## Abstract

**Background:** Large middle cerebral artery (MCA) infarctions can result in life-threatening cerebral edema. Quantitative brain atrophy may improve risk stratification for severe edema. We examined whether quantitative brain atrophy is associated with severe midline shift after large ischemic stroke and whether incorporating atrophy improves prediction beyond established clinical and radiographic predictors.

**Methods:** This was a retrospective observational cohort study of patients with ≥½ MCA ischemic infarction, presentation within 24 hours of last known well, and at least one follow-up head CT, admitted to two academic hospitals with comprehensive stroke centers between 2006 and 2024. The study was approved by the institutional review boards of both centers. Brain atrophy was quantified as the inverse of standardized brain volume on admission head CT. The primary outcome was severe radiographic mass effect, defined as midline shift ≥5 mm on follow-up CT. The secondary outcome was in-hospital mortality. Multivariable regression models assessed associations between quantified atrophy and outcomes. Incremental prognostic value was evaluated by comparing models with and without atrophy using measures of goodness of fit, calibration, and discrimination.

**Results:** Among 565 patients (mean age 67.5±15.7 years; 49.9% female), 223 (39.5%) developed severe mass effect. Greater atrophy was associated with lower odds of midline shift ≥5 mm (OR 0.44, 95% CI 0.34-0.58), but not with in-hospital mortality. Incorporation of atrophy significantly improved prediction of severe mass effect compared to the baseline model (likelihood ratio test χ² (1) = 41, p <0.001; AIC 703 vs. 741; BIC 733 vs. 767; AUC 0.68 vs. 0.60).

**Conclusions:** Quantified brain atrophy is independently associated with a reduced risk of severe mass effect after large MCA stroke and improved the performance of established predictive models. Incorporation of this imaging biomarker may enhance early risk stratification, monitoring, and intervention planning for patients at risk of life-threatening cerebral edema.

## INTRODUCTION

Malignant cerebral edema and resultant mass effect is a devastating and feared consequence of large middle cerebral artery (MCA) stroke, leading to high morbidity and mortality.^1–4^ Patients are at highest risk within the first week of stroke;^5–7^ however the timing, rate, and severity of brain swelling with or without concurrent hemorrhage remain unpredictable.^8–10^

Following ischemic injury, energy failure leads to ionic pump dysfunction, which causes intracellular water accumulation and widespread brain swelling.^11^ This rapid expansion generates a space occupying mass effect, which can elevate intracranial pressure, produce midline shift, precipitating herniation, neurologic deterioration and potentially death.^2,12^ Therefore, early identification of patients who may develop post-stroke malignant cerebral edema is critical,^3^ as it may allow timely interventions such as decompressive hemicraniectomy^13–15^ or hypothermia^16,17^ to alleviate neurologic deterioration.

Common models that currently serve as proxy quantitative tools for predicting neurologic deterioration include the EDEMA score,^18^ DASH score,^19^ and ACORNS scale after endovascular thrombectomy.^20^ While few small studies suggest that decreased brain volume, or greater brain atrophy, confers protection against malignant cerebral edema,^21,22^ no large-scale studies have validated a linear dose-response relationship, in part due to the lack of quantitative brain volumetrics. Recent advances in automated brain and cerebrospinal fluid (CSF) and brain parenchyma segmentation enable quantitative measurements of brain volume in larger cohorts,^23–27^ enabling a quantitative assessment of atrophy, defined as the inverse of standardized brain volume (total brain volume divided by intracranial volume). Our a priori hypothesis was that increased quantitative atrophy is associated with lower incidence of severe mass effect. We did not anticipate that atrophy would be associated with lower mortality, given known associations between age, dementia and atrophy with mortality.^28–31^

In this study, we examined whether greater quantitative brain atrophy is associated with a reduced risk of severe radiographic mass effect (≥5 mm midline shift)^32–34^ after large ischemic stroke (our primary hypothesis) and explored whether incorporating atrophy improves prediction of clinically relevant outcomes, including midline shift and in-hospital mortality.

Understanding the relationship between quantitative atrophy and mass effect severity in ischemic stroke could help refine earlier identification of patients at high risk for deterioration following large cerebral artery stroke, potentially guiding timely monitoring and intervention.

## METHODS

### Study Design and Participants

We conducted a retrospective cohort study of patients with a diagnosis of large (≥½-territory) MCA ischemic stroke, according to a previously published protocol, from Mass General Brigham (2006 to 2018) and Boston Medical Center (2019 to 2024).^35,36^ Eligible patients were admitted within 24 hours of last known well, and had available Digital Imaging and Communications in Medicine (DICOM) images that could be successfully segmented for volumetric analysis. Full eligibility criteria are listed in **Supplementary Figure S1**.

The study was approved by the Institutional Review Boards (IRB) at Mass General Brigham (2017P002564) and Boston Medical Center (H-37699). Informed consent was waived due to the retrospective design and use of standard-of-care procedures. All methods were performed in accordance with the relevant IRB guidelines and regulations. We reported the study adhering to the Strengthening the Reporting of Observational Studies in Epidemiology (STROBE) guidelines.^37^

### Data Collection and Measurement

Electronic medical record data included demographics, past medical history, admission stroke severity (National Institute of Health Stroke Scale (NIHSS)),^38^ last known well date and time, and modified Rankin Scale (mRS) score.^39^ We also collected data on administered procedures and treatments, including intravenous thrombolysis, mechanical thrombectomy, and decompressive hemicraniectomy. The earliest available head computed tomography (CT) image for each patient was obtained in DICOM format for analysis. Imaging characteristics including the Alberta Stroke Program Early CT Score (ASPECTS),^40^ MCA involvement, midline shift (measured at the level of the septum pellucidum) and presence/severity of hemorrhagic transformation^41^ were collected by trained team members (SC, LA) who were blinded to patient clinical data and quantitative atrophy measurements. Inter-rater reliability was established for all variables and was defined as >80% on at least 10% of the data. Instances where there was a discrepancy underwent third independent review (CJO) and were discussed to reach a consensus. Further details on data collection and ascertainment are outlined in **Supplementary Methods**.

### Exposures

The primary exposure was continuous quantitative brain atrophy, defined as the inverse of standardized brain volume. Standardized brain volume was derived from admission non-contrast head CT images. Imaging preprocessing included skull stripping and tissue segmentation into parenchyma, cerebrospinal fluid, and infarct-related hypodensity using a U-Net–based deep learning model.^24–27^ Detailed imaging methods are provided in the Supplementary Methods. To account for skull size and sex-based differences, absolute brain parenchymal volume was normalized by total intracranial volume to derive standardized brain volume (lower values indicate greater atrophy).

In exploratory analyses, we also calculated relative brain age, a measure capturing how “old” or “young” a brain appears compared to chronological age.^42^ Predicted brain age was first estimated from standardized brain volume, and age-related bias was then removed using a previously validated procedure. Relative brain age was defined as the residual difference between predicted and expected brain age for a given chronological age, where positive values indicate a structurally older-appearing brain whereas negative values represent a “younger-appearing” brain compared with age-matched peers. Detailed methods are provided in the **Supplementary Methods**.

### Outcomes

The primary outcome was severe radiographic mass effect, defined as midline shift ≥5 mm, a standard radiographic marker of malignant cerebral edema.^18,35,43–47^ The secondary outcome was in-hospital mortality. Exploratory outcomes included alternative midline shift thresholds associated with poor outcome (≥3 mm^48^ and ≥8 mm)^35,48^, to capture a broader spectrum of edema severity described in prior studies. Further exploratory outcomes are included in **Supplementary Methods.**

We explored additional clinical outcomes including potentially lethal malignant edema, defined as death due to cerebral edema (with midline shift ≥5 mm) or the need for decompressive hemicraniectomy as has been done in prior studies,^18^ and poor functional outcome at discharge. Poor functional outcome was defined using two thresholds, mRS score ≥4 (moderately severe disability, severe disability, or death) and mRS ≥5 (severe disability or death), to reflect differing patient and clinician perspectives on what constitutes an unacceptable degree of disability.^49,50^ Full variable definitions are provided in **Supplementary Methods.**

### Covariates

Covariates were selected a priori based on established predictors from the validated EDEMA model^18^ to create a parsimonious, clinically relevant model. These included chronological age, baseline hyperglycemia (glucose ≥150 mg/dL), National Institutes of Health Stroke Scale (NIHSS) score, acute reperfusion therapy (intravenous thrombolysis or mechanical thrombectomy), and early infarct volume (maximum infarct volume measured on head CT prior to outcome occurrence).

### Missing Data

For baseline variables with missing admission values but with longitudinal measurements available within 10 days of last known well, the earliest value within this window was carried backward and used as the baseline value. For all other baseline covariates, missingness was minimal among variables included in the primary analysis (<10%; Table S1). To retain all eligible participants, we applied a pragmatic single-imputation strategy. Specifically, missing values in continuous covariates were imputed using the cohort mean, and missing values in binary covariates were imputed to the majority category (i.e., assigned 0 when the observed proportion was ≤0.5 and 1 otherwise). As a sensitivity analysis, we repeated the primary analyses using multiple imputation by chained equations with predictive mean matching (MICE-PMM) for covariates with ≥5% missingness.

### Descriptive and Correlation Analyses

We summarized patient baseline characteristics as mean ± standard deviation (SD) or median with interquartile range (IQR) for continuous variables, and as counts with percentages for categorical variables. We compared patients with and without severe radiographic mass effect using t-tests or Wilcoxon rank-sum tests for continuous variables, and χ² or Fisher’s exact tests for categorical variables, as appropriate. We assessed correlations between quantitative atrophy and continuous measures using Spearman’s rank correlation coefficient and displayed the results in a correlation heatmap. Raincloud plots were used to display differences across dichotomous clinical outcomes and 10-year age strata, and midline shift distributions were plotted by quartiles of atrophy.

### Outcome Modeling

To test our primary hypothesis that increased quantitative atrophy was associated with lower severe mass effect, we used multivariable regression, adjusting for covariates. In subgroup analyses, we excluded patients with parenchymal hemorrhage type 2 defined by ECASS criteria (hematoma >30% of infarct zone),^41^ and subgroups stratified by hospital. To address potential differences in practice over time, we conducted a subgroup analysis of patients pre and post 2015. Adjusted effect sizes with 95% confidence intervals (CI) stratified by age groups were summarized in forest plots.

To assess the incremental predictive value of atrophy for clinical outcomes, we compared a base model containing the quantitative predictors from the EDEMA model (baseline hyperglycemia (glucose ≥150 mg/dL), National Institutes of Health Stroke Scale (NIHSS) score, acute reperfusion therapy (intravenous thrombolysis or mechanical thrombectomy), and early infarct volume (maximum measured on head CT prior to outcome occurrence) with an extended model that additionally included atrophy.

We reported standardized odds ratios (OR) as effect size per SD and CI for each predictor. We assessed model performance using goodness of fit, discrimination, and calibration/accuracy. We assessed model fit with the likelihood ratio test (LRT),^51^ Akaike Information Criterion (AIC),^52^ and Bayesian Information Criterion (BIC).^52^ For binary outcomes, we reported discrimination using area under the receiver operating characteristic curve (AUC),^53^ calibration and predictive accuracy using the Brier score,^53^ mean absolute error derived from calibration plots, and bias-corrected calibration curves.^54^ For continuous outcomes, we reported explanatory power through adjusted R², and predictive accuracy using root mean squared error.^55^

To reduce optimism and enhance generalizability, AUC, Brier score, and root mean squared error were averaged over 5-fold cross-validation,^56^ while calibration plots and mean absolute error were bias-corrected by bootstrap resampling (200 iterations).^57^ All statistical analyses were conducted using R software (Version 4.5.0), with two-sided p<0.05 considered statistically significant for our primary hypothesis. Other analyses are hypothesis generating.

## RESULTS

### Study Cohort and Baseline Characteristics

A total of 565 patients met inclusion criteria (**Supplementary Figure 1**). The mean age of the cohort was 67.5 ± 15.7 years; 282 (49.9%) were female. The median time from the last known well to head CT was 11.5 hours [IQR 5.0-23.2]. Two hundred and twenty-three (39.5%) developed severe mass effect.

Patients with severe mass effect were younger, had lower ASPECTS scores, and higher incidence of hemorrhagic transformation (76.7% vs. 45.6%) particularly parenchymal hemorrhage type 2 (13.5% vs 1.5%). They also had larger early infarct volumes (which may not reflect ultimate injury volume) (96.7 [41.4-152.8] mL vs. 86.7 [25.2-138.6] mL), higher in-hospital mortality (39.9% vs. 20.2%), and less atrophy (1.09 [IQR 1.06-1.12] vs. 1.13 [IQR 1.08-1.17]). Details are provided in **Table 1** and missing Data summary in **Supplementary Table S1**.

**Table 1.**
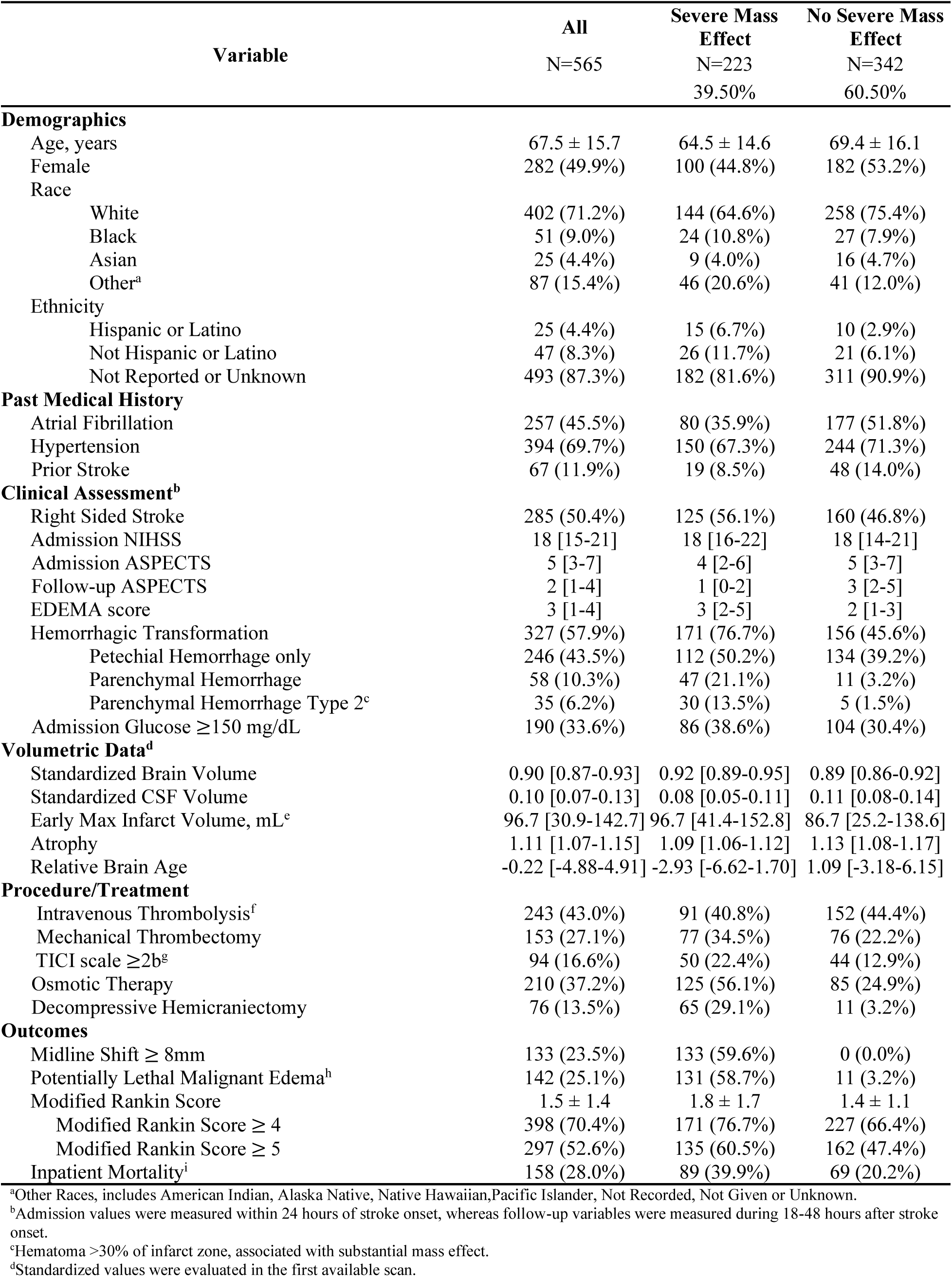
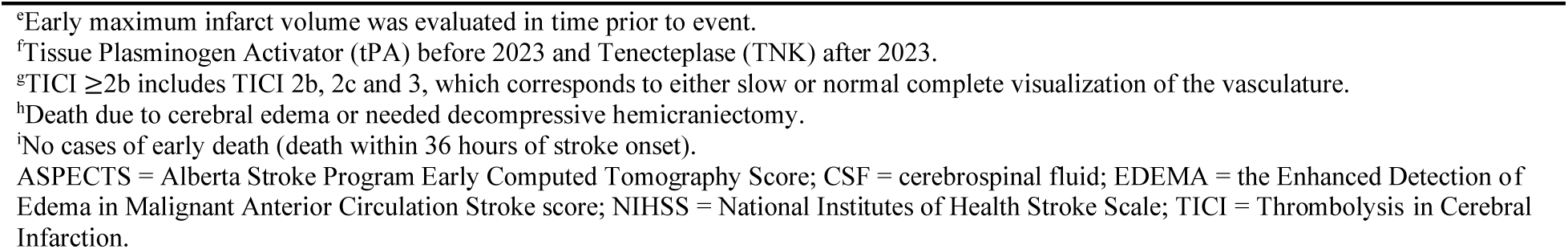
Patient Characteristics.

| Variable | All<br>N=565 | Severe Mass<br>Effect<br>N=223<br>39.50% | No Severe Mass<br>Effect<br>N=342<br>60.50% |
| --- | --- | --- | --- |
| **Demographics** |  |  |  |
| Age, years | 67.5 ± 15.7 | 64.5 ± 14.6 | 69.4 ± 16.1 |
| Female | 282 (49.9%) | 100 (44.8%) | 182 (53.2%) |
| Race |  |  |  |
| White | 402 (71.2%) | 144 (64.6%) | 258 (75.4%) |
| Black | 51 (9.0%) | 24 (10.8%) | 27 (7.9%) |
| Asian | 25 (4.4%) | 9 (4.0%) | 16 (4.7%) |
| Other<sup>a</sup> | 87 (15.4%) | 46 (20.6%) | 41 (12.0%) |
| Ethnicity |  |  |  |
| Hispanic or Latino | 25 (4.4%) | 15 (6.7%) | 10 (2.9%) |
| Not Hispanic or Latino | 47 (8.3%) | 26 (11.7%) | 21 (6.1%) |
| Not Reported or Unknown | 493 (87.3%) | 182 (81.6%) | 311 (90.9%) |
| **Past Medical History** |  |  |  |
| Atrial Fibrillation | 257 (45.5%) | 80 (35.9%) | 177 (51.8%) |
| Hypertension | 394 (69.7%) | 150 (67.3%) | 244 (71.3%) |
| Prior Stroke | 67 (11.9%) | 19 (8.5%) | 48 (14.0%) |
| **Clinical Assessment<sup>b</sup>** |  |  |  |
| Right Sided Stroke | 285 (50.4%) | 125 (56.1%) | 160 (46.8%) |
| Admission NIHSS | 18 [15-21] | 18 [16-22] | 18 [14-21] |
| Admission ASPECTS | 5 [3-7] | 4 [2-6] | 5 [3-7] |
| Follow-up ASPECTS | 2 [1-4] | 1 [0-2] | 3 [2-5] |
| EDEMA score | 3 [1-4] | 3 [2-5] | 2 [1-3] |
| Hemorrhagic Transformation | 327 (57.9%) | 171 (76.7%) | 156 (45.6%) |
| Petechial Hemorrhage only | 246 (43.5%) | 112 (50.2%) | 134 (39.2%) |
| Parenchymal Hemorrhage | 58 (10.3%) | 47 (21.1%) | 11 (3.2%) |
| Parenchymal Hemorrhage Type 2<sup>c</sup> | 35 (6.2%) | 30 (13.5%) | 5 (1.5%) |
| Admission Glucose ≥150 mg/dL | 190 (33.6%) | 86 (38.6%) | 104 (30.4%) |
| **Volumetric Data<sup>d</sup>** |  |  |  |
| Standardized Brain Volume | 0.90 [0.87-0.93] | 0.92 [0.89-0.95] | 0.89 [0.86-0.92] |
| Standardized CSF Volume | 0.10 [0.07-0.13] | 0.08 [0.05-0.11] | 0.11 [0.08-0.14] |
| Early Max Infarct Volume, mL<sup>c</sup> | 96.7 [30.9-142.7] | 96.7 [41.4-152.8] | 86.7 [25.2-138.6] |
| Atrophy | 1.11 [1.07-1.15] | 1.09 [1.06-1.12] | 1.13 [1.08-1.17] |
| Relative Brain Age | -0.22 [-4.88-4.91] | -2.93 [-6.62-1.70] | 1.09 [-3.18-6.15] |
| **Procedure/Treatment** |  |  |  |
| Intravenous Thrombolysis<sup>f</sup> | 243 (43.0%) | 91 (40.8%) | 152 (44.4%) |
| Mechanical Thrombectomy | 153 (27.1%) | 77 (34.5%) | 76 (22.2%) |
| TICI scale ≥2b<sup>g</sup> | 94 (16.6%) | 50 (22.4%) | 44 (12.9%) |
| Osmotic Therapy | 210 (37.2%) | 125 (56.1%) | 85 (24.9%) |
| Decompressive Hemicraniectomy | 76 (13.5%) | 65 (29.1%) | 11 (3.2%) |
| **Outcomes** |  |  |  |
| Midline Shift ≥ 8mm | 133 (23.5%) | 133 (59.6%) | 0 (0.0%) |
| Potentially Lethal Malignant Edema<sup>h</sup> | 142 (25.1%) | 131 (58.7%) | 11 (3.2%) |
| Modified Rankin Score | 1.5 ± 1.4 | 1.8 ± 1.7 | 1.4 ± 1.1 |
| Modified Rankin Score ≥ 4 | 398 (70.4%) | 171 (76.7%) | 227 (66.4%) |
| Modified Rankin Score ≥ 5 | 297 (52.6%) | 135 (60.5%) | 162 (47.4%) |
| Inpatient Mortality<sup>i</sup> | 158 (28.0%) | 89 (39.9%) | 69 (20.2%) |

<sup>a</sup>Other Races, includes American Indian, Alaska Native, Native Hawaiian, Pacific Islander, Not Recorded, Not Given or Unknown.
<sup>b</sup>Admission values were measured within 24 hours of stroke onset, whereas follow-up variables were measured during 18-48 hours after stroke onset.
<sup>c</sup>Hematoma >30% of infarct zone, associated with substantial mass effect.
<sup>d</sup>Standardized values were evaluated in the first available scan.
<sup>e</sup>Early maximum infarct volume was evaluated in time prior to event.
<sup>f</sup>Tissue Plasminogen Activator (tPA) before 2023 and Tenecteplase (TNK) after 2023.
<sup>g</sup>TICI $\geq 2b$ includes TICI 2b, 2c and 3, which corresponds to either slow or normal complete visualization of the vasculature.
<sup>h</sup>Death due to cerebral edema or needed decompressive hemicraniectomy.
<sup>i</sup>No cases of early death (death within 36 hours of stroke onset).
ASPECTS = Alberta Stroke Program Early Computed Tomography Score; CSF = cerebrospinal fluid; EDEMA = the Enhanced Detection of Edema in Malignant Anterior Circulation Stroke score; NIHSS = National Institutes of Health Stroke Scale; TICI = Thrombolysis in Cerebral Infarction.

### Atrophy

Across the cohort, atrophy increased with chronological age (Spearman r_s_=0.66, p<0.001). No apparent differences in atrophy were observed across sex or in-hospital mortality (**Figure 1, Supplementary Figure S2)**. Greater quantitative atrophy was independently associated with lower odds for severe mass effect, when adjusted for EDEMA model covariates (OR 0.44, 95% CI 0.34-0.58, per 1 SD increase in atrophy) (**Table 2)**. Significance was consistent in the multiple-imputation sensitivity analysis. Increased atrophy was not associated with in-hospital mortality (OR 0.87, 95% CI 0.67-1.14 per 1 SD increase in atrophy) **(Supplementary Table S2).**

**Figure 1.**
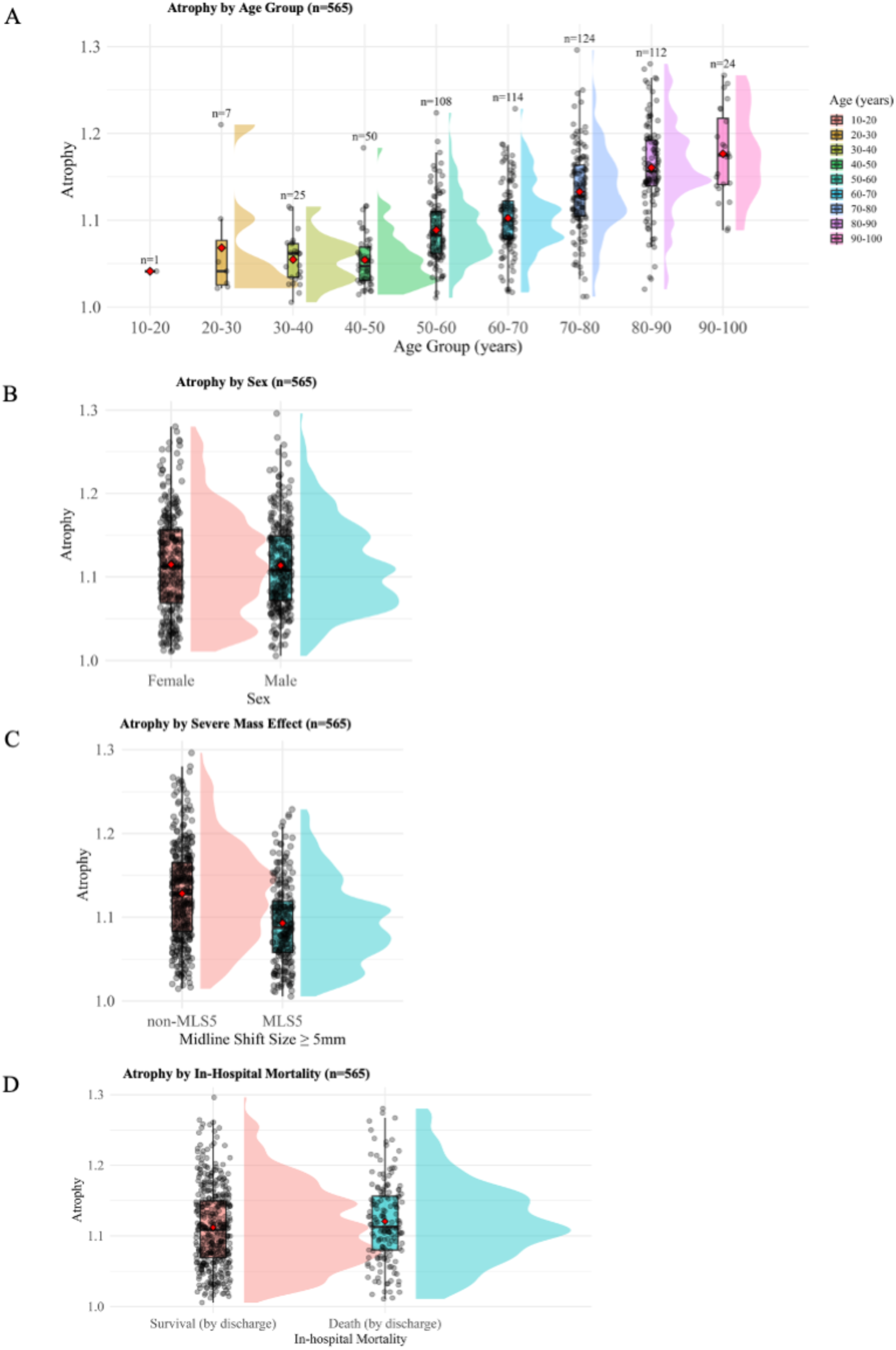
Atrophy stratified by age, sex, midline shift, and in-hospital mortality. Patients with (A) younger age and those with (C) severe mass effect tend to have lesser atrophy. No apparent differences in atrophy are observed across (B) sex or (D) in-hospital mortality.

**Table 2.** Logistic Regression of Brain Atrophy and Severe Mass Effect.

| Variables | Standardized Odds Ratio* |  |  | p Value |
| --- | --- | --- | --- | --- |
|  | Odds Ratio | 95% Confidence Intervals |  |  |
|  |  | Lower | Upper |  |
| Baseline Model |  |  |  |  |
| Age | 0.69 | 0.58 | 0.83 | <0.001 |
| NIHSS | 1.29 | 1.08 | 1.55 | 0.006 |
| Hyperglycemia | 1.51 | 1.04 | 2.19 | 0.030 |
| Intervention | 1.03 | 0.72 | 1.46 | 0.882 |
| Early Infarct Volume | 1.07 | 0.90 | 1.28 | 0.426 |
| Baseline Model with Atrophy |  |  |  |  |
| Age | 1.11 | 0.88 | 1.41 | 0.377 |
| NIHSS | 1.29 | 1.07 | 1.55 | 0.008 |
| Hyperglycemia | 1.45 | 0.98 | 2.13 | 0.061 |
| Intervention | 0.90 | 0.62 | 1.30 | 0.562 |
| Early Infarct Volume | 0.98 | 0.81 | 1.18 | 0.795 |
| Atrophy | 0.44 | 0.34 | 0.58 | <0.001 |

\* Standardized odds ratios quantified the effects of a 1 standard deviation increase in each continuous predictor. Standard deviation for each continuous predictors are: age, 15.7 years; NIHSS, 5.7; Early Infarct Volume, 71.1 mL; Atrophy, 0.06. NIHSS = National Institutes of Health Stroke Scale

Increased atrophy was associated with lower odds of midline shift ≥8 mm (OR 0.40, 95% CI 0.29-0.55; p<0.001), as well as lower potentially lethal malignant edema (OR 0.57, 95% CI 0.41-0.78; p<0.001). Sensitivity analyses excluding parenchymal hemorrhage type 2, the Mass General Brigham only, the pre-2015 or post-2015 cohorts produced results consistent with our primary analysis (**Supplementary Figures S3-5, Supplementary Tables S2-4).** Age-stratified models retained significance in both patients greater and less than 60 years of age **(Figure 2, Supplementary Table S5)**.

**Figure 2.**
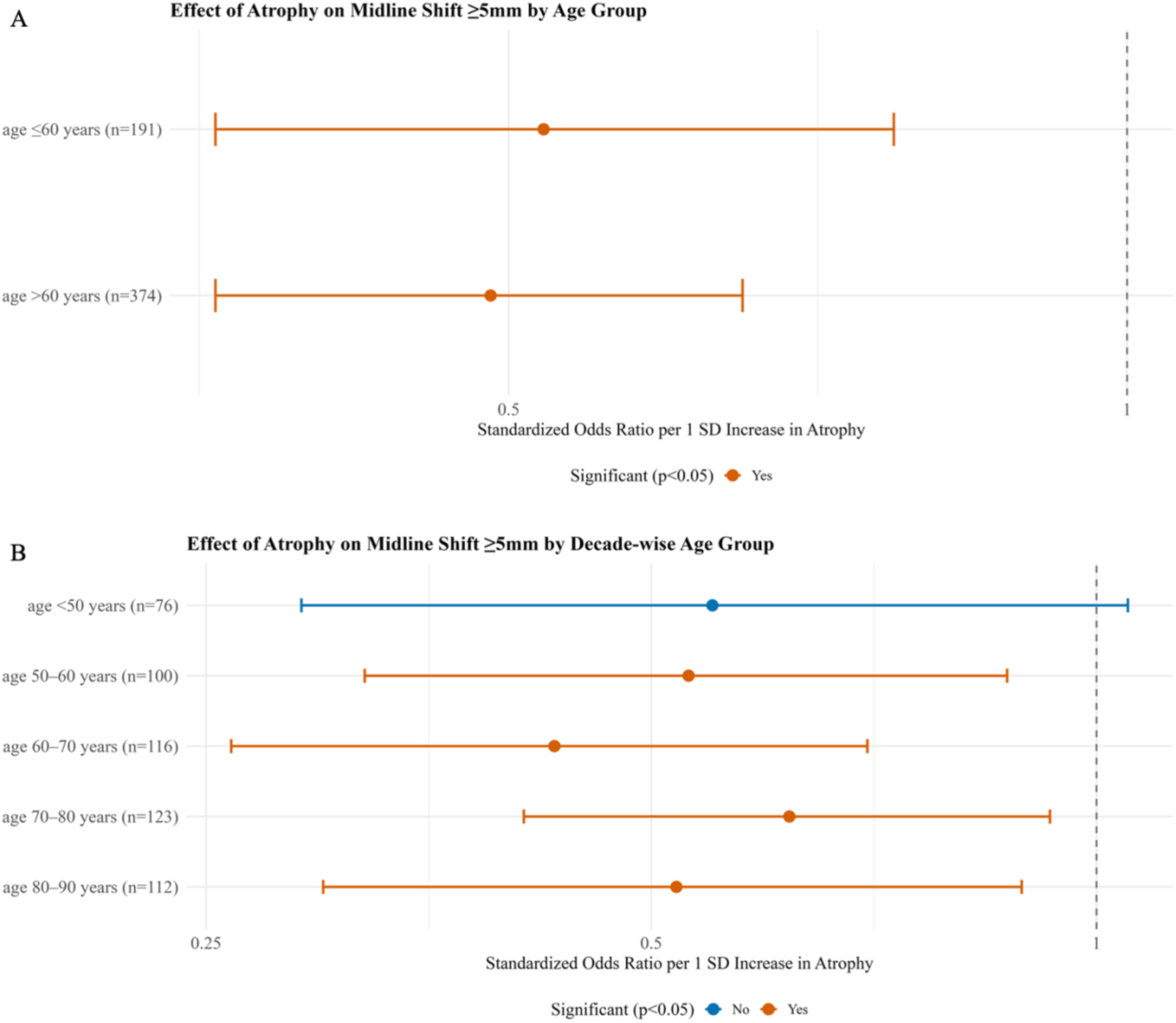
Effect size of atrophy on severe mass effect, (A) dichotomized into two age groups and (B) stratified into 10-year age groups. The protective effect of larger atrophy is significant only in patients older than 60 years. Error bars represent 95% confidence intervals. SD = standard deviation.

We observed similar findings between relative brain age and the primary outcome (OR 0.54, 95% CI 0.45-0.66, per 10-years) (**Supplementary Figure S6, Supplementary Table S6)**. The association between relative brain age and chronological age, as well as clinical outcomes, were similar to the findings with quantitative atrophy and included in **Supplementary Figures S7-8 and Supplementary Tables S7-9.**

### Model Performance Comparison

In the baseline logistic regression model, higher admission NIHSS and baseline hyperglycemia were significantly associated with higher odds of severe mass effect. Adding quantified atrophy in the baseline model significantly improved goodness of fit as reflected by the LRT, (LRT χ² (1) = 41; p <0.001) and better balance between goodness of fit and model complexity, reflected by lower AIC and BIC values (AIC 703 vs. 741; BIC 733 vs. 767), modestly higher discrimination (AUC 0.68 vs. 0.60), and improved calibration/accuracy (Brier score 0.22 vs. 0.23; Mean Absolute Error 0.01 vs. 0.03) (**Table 3, Supplementary Figure S9**). When stratified by admission era (pre-2015: n=331; post-2015: n=234), adding atrophy improved discrimination in both groups. In the pre-2015 cohort, AUC increased from 0.60 (baseline) to 0.67 (baseline + atrophy) and in the post-2015 cohort, AUC increased from 0.62 to 0.70 respectively (**Supplementary Table S10**).

**Table 3.**
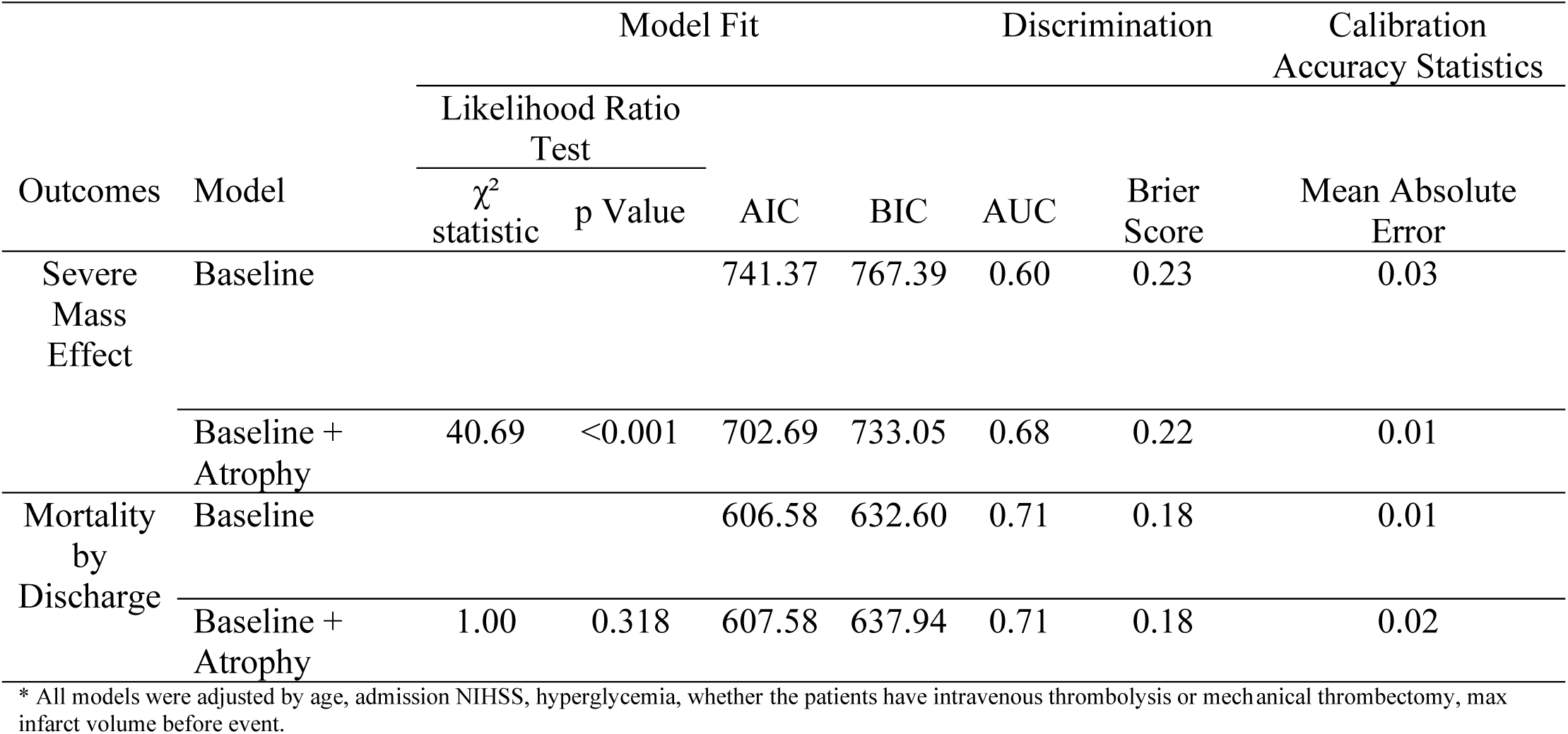
Comparison of Logistic Regression Models with and without Atrophy.

There was no improvement in predicting mortality (**Table 3, Supplementary Figure S10**). Models that included relative brain age demonstrated results consistent with brain atrophy (**Supplementary Figure S10-11, Supplementary Table S11,12)**.

## DISCUSSION

In this study, we observed that greater baseline brain atrophy was associated with a lower likelihood of developing severe mass effect after large MCA ischemic stroke. Our work confirms and builds upon anecdotal clinical observations that patients with substantial brain atrophy are less likely to develop malignant edema.^21,22^ This relationship appeared linear across atrophy quartiles, suggesting an effect dependent on the degree of quantified atrophy. Incorporating quantified atrophy into a baseline predictive model significantly improved the ability to identify patients at risk for severe mass effect, underscoring the relevance of baseline structural brain characteristics in determining edema susceptibility beyond established clinical and imaging factors.

Prior studies have proposed that brain atrophy may protect against malignant cerebral edema by providing intracranial volume reserve.^21,22^ However, these investigations were constrained by small sample sizes and relied on qualitative assessments. Our study advances this work by applying deep learning–based volumetric segmentation to generate continuous, standardized measures of intracranial volume and atrophy.^24–27^ We further observed that quantitative atrophy provides independent predictive value beyond established clinical and imaging factors included in the EDEMA score, improving model fit and discrimination and demonstrating that it is not merely colinear with existing predictors. When stratified by age, the inverse association between quantified atrophy and severe mass effect remained significant among patients aged ≥60 years but not among younger individuals, likely reflecting a broader range of atrophy values in the older population.

We propose that brain atrophy reduces susceptibility to mass effect through two complementary mechanisms. First, smaller parenchymal volume results in less tissue available to swell, thereby limiting total edema volume and its mechanical impact. Second, in patients with atrophy, the increased intracranial reserve allows for greater displacement of swollen tissue without compressing midline structures. A larger cerebrospinal fluid to brain volume ratio provides additional buffering capacity against rising intracranial pressure, mitigating midline shift.^21,58^

Interestingly, although increased atrophy appeared protective against severe mass effect, it was not associated with improved clinical outcomes, including mortality, need for decompressive hemicraniectomy, or favorable discharge mRS. This suggests that while atrophy may provide a mechanical buffer against edema-related midline shift, it does not necessarily confer protection from poor neurological or functional recovery. One explanation may be that atrophy reflects an underlying vulnerability and reduced brain reserve, limiting recovery potential even in the absence of malignant edema. Furthermore, surgical decisions and mortality risk are often influenced by age and comorbidities, which may attenuate the apparent protective effect of atrophy.

Despite this, our findings remain clinically relevant. Patients with increased atrophy may be at lower risk for life-threatening mass effect and could benefit from tailored management strategies such as less time undergoing hourly neurological assessments which can reduce sleep and increase delirium,^59,60^ prioritization of medical rather than surgical therapy, and consideration for step-down or non-ICU care to reduce secondary complications. Avoiding unnecessary ICU exposure may be particularly important in older patients or those with vascular cognitive impairment, since delirium and prolonged hospitalization are known to worsen long-term outcomes.^61^

We recognize several limitations. The dual-center observational cohort design may limit the generalizability of our findings to broader populations or other clinical settings. Although we adjusted for hypothesized confounding factors through statistical methods, residual confounding may still be present due to unmeasured or unknown variables. Imaging was obtained at clinician discretion, which may lead to ascertainment bias. Other clinical factors, including timing of medical management including osmotic medications, and patient-specific comorbidities, may also have influenced observed outcomes. We did not have information on important relevant outcomes including neurologic deterioration. We also did not have information on potential other factors associated with edema progression including collateral score, success of reperfusion therapy, and CT perfusion.^20,62–64^ Withdrawal of life-sustaining therapy and its age-related bias, may also influence measured outcomes.^65,66^ While these factors may influence edema development, our primary analyses incorporated the validated EDEMA score, which provides a parsimonious and clinically relevant framework for risk stratification. In addition, subgroup analyses suggested that the predictive value of atrophy for severe edema is strongest in patients over 60, limiting the generalizability of this finding across all ages. Withdrawal of life-sustaining therapies likely impacts older patients more than younger ones, which could negate any protective effect of brain atrophy or reduced mortality after stroke. Future studies should validate these findings prospectively in multi-center cohorts and explore multimodal brain age models incorporating structural, functional, and molecular imaging features.

## CONCLUSION

Increased quantitative atrophy is associated with lower likelihood of severe mass effect in patients with large middle cerebral artery stroke, highlighting the potential benefit of standardizing how quantitative brain atrophy is used in risk prediction and triaging of malignant edema.

## Acknowledgements

CJO receives support from NIH/NINDS K23NS116033; American Heart Association 23CDA1041762. RD receives support from NIH R01NS1121218 and U24NS132940

## Author Contributions

CJO, RD, and YD contributed to conception and design of the study. CJO, SC, RS, LAM, and MKA contributed to data acquisition including manually reviews of patient notes and radiographic images to determine radiographic outcomes. YD and NJI contributed to statistical analysis. YD, CJO, AZA, LAM and NJI contributed to interpretation of results and preparing figures. YD, AZA, and CJO contributed to drafting the text. AK, AA, MKA, RD contributed to image analysis and interpretation. YD, AZA, NJI, SC, LAM, ALR, RS, AK, AA, MKA, HC, DMG, RD, and CJO contributed to critical editing and revision. CJO provided overall study direction and critical review. All authors approved the submitted version.

## Potential Conflicts of Interest

Nothing to report.

## Data Availability

Anonymized data that support the findings of this study are available from the corresponding author upon request with data use agreements.

---

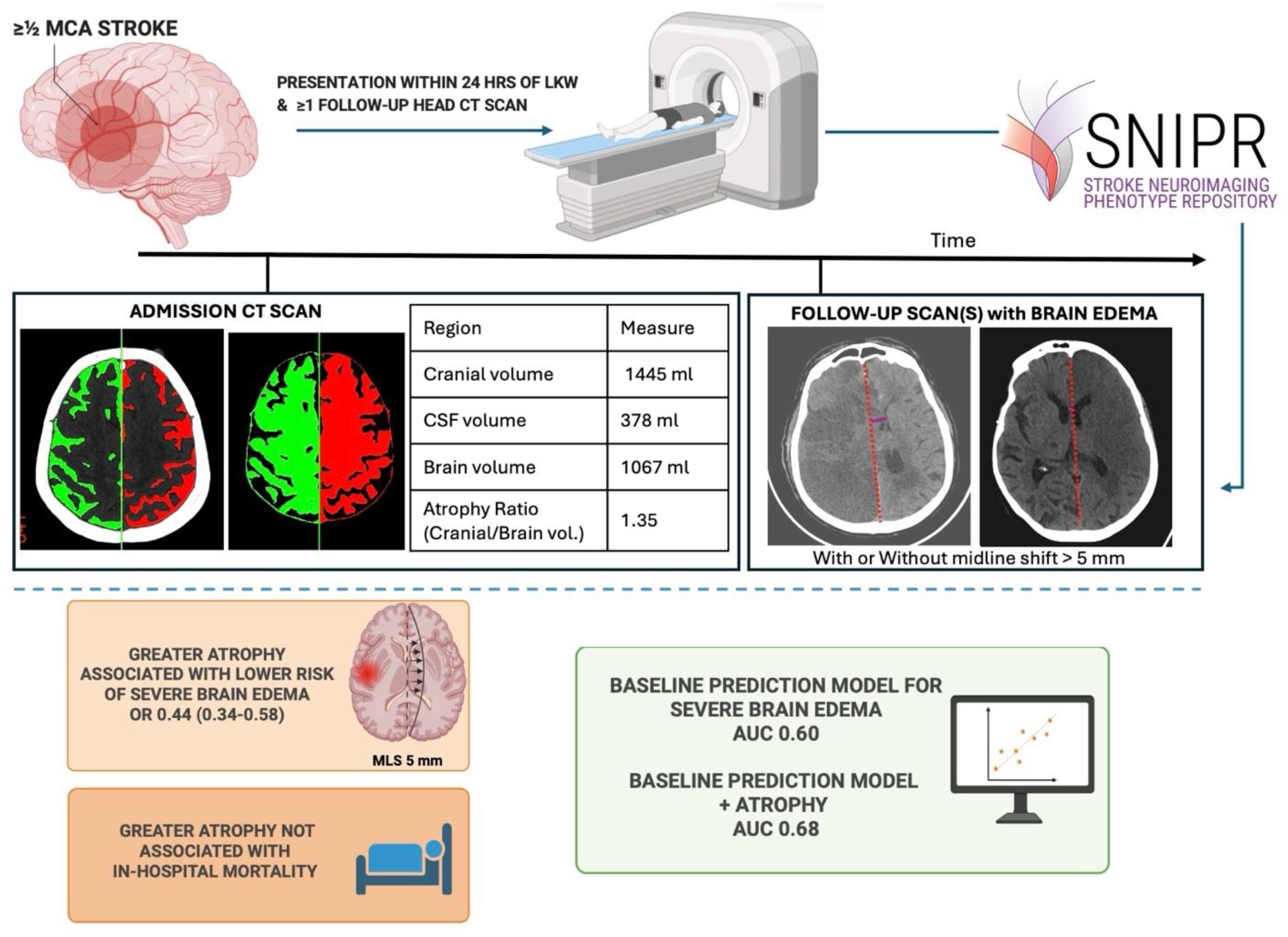

## Notes

### Competing Interest Statement

The authors have declared no competing interest.

### Author Declarations

The study was approved by the Institutional Review Boards (IRB) at Mass General Brigham (2017P002564) and Boston Medical Center (H-37699).

