## Supplementary File for "Quantified Brain Atrophy and Risk of Severe Mass Effect in Acute Ischemic Stroke"

### Supplementary Methods

#### Data Collection and Ascertainment

Demographics, clinical variables including past medical history, stroke assessment, procedures and treatment, and outcomes, were extracted from the electronic medical records at Boston Medical Center and Mass General Brigham. We included the following ascertainment rules that pertain to relevant variables.

*Middle Cerebral Artery (MCA) involvement criteria* were adjudicated by the principal investigator (CJO) as ≥1/2 MCA territory either on Computed Tomography (CT) or Magnetic Resonance Imaging (MRI) scan during admission by visualization of either full involvement of the superior or inferior division of the MCA territory or sufficient hypodensity/diffusion restriction of both superior and inferior divisions to equate to 50 or more percent of the full MCA territory. Among patients who had available volumetric imaging on CT (N=58), median infarct volume was 210.7 cc (IQR 124.6-246.2 cc).

*Last known well* date times were identified via clinical notes. Last known well was considered the last time before a patient had a significant deficit as inferred by National Institutes of Health Stroke Score (**NIHSS**) (times in which symptoms of numbness, tingling, or other symptoms that would result in NIHSS<3 were not taken as the time of last known well). Last known well without exact times or only reference to time were labeled as "inferred" (N=1). If last known well referred to "last night before bed" with no exact time, then last known well was assumed to be 10:00 PM. All last known well data underwent supervisor review (CJO).

**Imaging related variables** including admission Alberta Stroke Program Early CT Score (ASPECTS), hemorrhagic transformation, and midline shift at the septum pellucidum were determined by trained team members (SC, LA) after establishing satisfactory inter-rater reliability defined as >80% of 10-15% of the data (please see below for precise sample size). Instances in which there was a discrepancy regarding radiographic features underwent a third review (CJO) and were discussed to reach consensus.

- The *ASPECTS* divides the brain parenchyma into 10 separate non-overlapping regions, seven at the level of the thalamus and basal ganglia, and three at the level just rostral to the ganglionic structures. For each baseline non-contrast CT scan (NCCT), a region was considered affected if there was radiographic evidence of ischemia, including hypodensity, loss of cortical ribbon, and sulcal effacement.[4] To be counted as affected, the lesion had to involve at least 10% of the respective region. An ASPECTS score of 10 indicates that all examined regions at the two assessed levels are normal, whereas a score of 0 indicates that all predetermined regions are affected by ischemia. A review of 14% of scans resulted in a percent agreement between the two reviewers of 90% for dichotomous ASPECTS categories (10-8, 7-0), and 80% for high, medium, and low ASPECTS (10-8, 7-4, 3-0).
- *Hemorrhagic transformation* was recorded by manual inspection according to European Cooperative Acute Stroke Study (ECASS) II criteria.[5] The classifications included hemorrhagic infarction, parenchymal hemorrhage, and subarachnoid hemorrhage. Petechial hemorrhages alone were classified as Petechial hemorrhage 1 (HI1) and petechial hemorrhages starting to gain mild confluence but without any mass effect were

classified as Petechial hemorrhage 2 (HI2). Hemorrhage involving ≤30% of the infarcted area or causing minor mass effect attributable to hematoma was classified as parenchymal hemorrhage 1 (PH1). Hemorrhage involving > 30% of the infarcted zone or causing substantial mass effect attributable to hematoma was classified to be parenchymal hemorrhage 2 (PH2). Percent agreement between trained team members (SC, CJO) on a 14% sample was 80%.

- *Midline shift* was measured at the level of the septum pellucidum by a trained member of the team using imaging viewer software Centricity™ Enterprise Web v.3.0 (GE healthcare, US) by navigating to the slice demonstrating maximum midline shift. The reviewer drew a line connecting the anterior attachment of the falx cerebri (the most frontal notch visible on the scan) and the occipital protuberance (the most posterior notch seen on the skull image). Windows were set at W:30, L:30. The perpendicular distance between the midline and the septum pellucidum was measured at both the most lateral and medial boundaries and the average of these two measurements was recorded as the final midline shift for analysis. The results were recorded in a CSV file and the difference between the manually measured midline shift and the midline shift reported in radiology reports were calculated. A blinded review of >10% of the scans showed a mean error of 1.19 mm between the two measurement methods.

#### Exposures

##### Brain Atrophy

Our primary exposure was *brain atrophy*, quantified as the inverse of standardized brain volume. Cerebrospinal fluid (CSF) volumetrics were derived from each CT scan using the deep learning pipeline detailed below.[1–4]

1. Preprocessing and Skull-Stripping

   Non-contrast head CT scans were converted to NIfTI format and preprocessed to isolate intracranial brain tissue. Skull-stripping was performed in two stages: an initial head extraction using level-set segmentation via the Computational Morphometry Toolkit (CMTK), followed by automated brain extraction using the FSL Brain Extraction Tool (BET). Intensity normalization was achieved by subtracting the mean brain intensity and dividing by twice the standard deviation.

2. Midline Estimation and Registration

   A CT template with pre-annotated midline landmarks was resampled and registered to each subject scan using affine transformation via FSL's FLIRT tool (12 degrees of freedom, correlation ratio as the cost function). The resulting transformation matrix was applied to the midline mask to align it with the subject space. For increased robustness, the midline coordinates were smoothed across slices using RANSAC-based linear regression.

3. Infarct Segmentation (Follow-up and Visible Baseline CTs)

   Regions of infarction were segmented using a deep learning model based on the U-Net architecture, trained to detect hypodense infarct regions on non-contrast CT images. The model produced a probabilistic infarct mask, which was thresholded to yield a binary segmentation. To ensure anatomical validity, the mask was post-processed using 3D region-growing, retaining only the largest connected component for subsequent analysis.

4. Cerebrospinal Fluid (CSF) Segmentation

CSF regions were segmented using a previously validated[2] U-Net–based deep learning model trained specifically for stroke-related CT scans. The resulting CSF mask was employed to exclude non-parenchymal voxels from normalized water uptake (NWU) calculations.

5. Hemispheric Division and Mask Mirroring

The transformed midline was used to bisect the brain into left and right hemispheres. Infarct masks were mirrored to the contralateral hemisphere using a series of geometric transformations—translation, rotation, reflection, and inverse affine registration—implemented via OpenCV. Voxels corresponding to CSF or falling outside predefined Hounsfield Unit (HU) thresholds (0–40 HU for infarct, 20–80 HU for normal brain tissue) were excluded prior to density analysis.

**Relative Brain Age**

In addition to atrophy we also explored *relative brain age*, a measure capturing how "old" or "young" a brain appears compared to chronological age. First, we estimated predicted brain age for each patient by fitting a linear regression model in which chronological age was predicted from standardized brain volume. Because predicted brain age is inherently correlated with chronological age, we applied an established procedure to remove this dependence and mitigate regression dilution bias.[5] To do this, we regressed predicted brain age on chronological age across the cohort to obtain expected brain age for a given chronological age. Relative brain age was defined as the residual difference between a patient's predicted brain age and expected brain age:

Brain Age = Predicted Age from Standardized Brain Volume

Expected Brain Age = Expected (Predicted Age | Chronological Age)

Relative Brain Age= Brain age – Expected Brain Age

Positive relative brain age values indicate a structurally "older-appearing" brain with greater atrophy for a given chronological age, whereas negative values represent a "younger-appearing" brain compared with age-matched peers.

#### Outcomes

The primary outcome was severe mass effect, defined as midline shift ≥5 mm, chosen as the primary radiographic marker of malignant cerebral edema in accordance with prior studies and guideline definitions.[6–12] The secondary outcome was inpatient mortality, defined as all-cause in-hospital mortality. Exploratory outcomes included additional measurements.

- We examined the *alternative definitions of midline shift*, including thresholds of midline shift ≥3 mm[13] and midline shift ≥8 mm,[10,13] previously used to indicate clinically significant mass effect and poor prognosis. Midline shift was also measured as a continuous variable, including the maximum value measured over the study period (within 10 days of last known well), and the early maximum value measured within 24 hours of last known well.

- *Potentially lethal malignant edema (PLME)* was defined as death due to cerebral edema in the presence of midline shift ≥5 mm or the need for decompressive hemicraniectomy (DHC), following previously established criteria.[6] We also included *DHC* alone as an outcome, defined as the performance of hemicraniectomy during hospitalization regardless of survival. Images collected after DHC were removed.
- *Functional outcomes at discharge* were assessed using the modified Rankin Scale (mRS), with poor outcome defined as either mRS ≥4 (moderately severe disability or worse) or mRS ≥5 (severe disability or worse), consistent with prior stroke outcome literature.[14,15]

#### Descriptive and Correlation Analyses

For atrophy and relative brain age, multivariable regression models were used to assess the incremental predictive value of atrophy or relative brain age beyond established clinical and imaging variables. Model performance was evaluated by measures of goodness of fit, discrimination, and calibration. To visualize group differences, we generated raincloud plots comparing atrophy and relative brain age across dichotomous clinical outcomes and across 10-year age groups. We also plotted the distribution of midline shift across quartiles of atrophy and relative brain age.

Calibration plots were not generated for continuous outcomes, as ordinary least squares estimation is unbiased by design and error metrics (adjusted $R^2$ and root mean squared error) already summarize model agreement.

**Pre vs Post 2015 Analysis**

We conducted a sensitivity analysis to evaluate secular trends in stroke care over the study period. Patients were classified by treatment era as pre-2015 vs post-2015 based on last known well or, if unavailable, presentation date. We refitted the primary MLS ≥5 mm model in the stratified groups to evaluate whether the association results and model performances differed across eras.

#### Supplementary Results

##### Study Cohort and Baseline Characteristics

A total of 565 patients met inclusion criteria (**Supplementary Figure 1**). Patients with severe mass effect had less atrophy (1.09 [IQR 1.06-1.12] vs. 1.13 [IQR 1.08-1.17]) and younger relative brain age (-2.93 [-6.62-1.70] vs. 1.09 [-3.18-6.15]). Details are included in **Table 1** and Missing Data in **Supplementary Table S1**.

##### Atrophy and Relative Brain Age

*Atrophy*

Across the cohort, atrophy increased with chronological age (Spearman $r_s$=0.66, p<0.001) (**Supplementary Figure S2**). Although atrophy was strongly associated with severe mass effect, it did not appear to be associated with our secondary outcome of in-hospital mortality, or any of the exploratory outcomes except midline shift ≥8 mm, and a trend towards significance with PLME (**Supplementary Table S3).** There were no apparent differences in atrophy between patients with and without modified Rankin Score ≥4 and modified Rankin Score ≥5 (**Supplementary Figure S3).** Atrophy correlated strongly with relative brain age ($r_s$ = 0.76, p<0.001) (**Supplementary Figure S4**). In quartile analyses, lower brain atrophy was associated with greater maximum midline shift within 10 days of last known well (**Supplementary Figure**

**S5, Supplementary Tables S3-S4).** Age-stratified models retained significance in patients both <60 and ≥60 years of age, especially those aged 60–70 years **(Supplementary Table S5)**

*Relative Brain Age*

Greater relative brain age was consistently associated with lower maximum midline shift within 10 days and lower odds of severe midline shift (≥8 mm) (**Supplementary Figure S5**). Greater relative brain age was associated with lower odds of severe mass effect (**Supplementary Table S6).** We did not observe significant associations with early (<24 hours) or mild (≥3 mm) midline shift. Notably, relative brain age was not associated with decompressive hemicraniectomy, poor discharge functional outcome, or in-hospital mortality **(Supplementary Figure S6).** The association of relative brain age with exploratory outcomes is displayed in **Supplementary Table S7.** Age-stratified models retained significance in patients both <60 and ≥60 years, especially those aged 60–70 years **(Supplementary Figure S7, Supplementary Tables S7 and S8)**. Across the cohort, relative brain age was independent of chronological age ($r_s = 0.07$, $p = 0.12$) (**Supplementary Figure S8)**. Sensitivity analyses excluding parenchymal hemorrhage type 2 and the Mass General Brigham only cohort produced consistent results. Full results are reported in **Supplementary Tables S4-S6**.

#### Multivariable Associations

In the baseline logistic regression model, higher admission NIHSS and baseline hyperglycemia were significantly associated with higher odds of severe mass effect. Adding relative brain age significantly improved the prediction of severe mass effect. Each 1-SD increase in relative brain age (i.e., older-appearing brain than chronological age) was associated with reduced odds of severe mass effect (OR 0.54, 95% CI 0.45–0.66, per SD 7.7 years; OR 0.42, 95% CI 0.24–0.71, per 10-year). Complete results are in **Supplementary Table S6.** Because atrophy and relative brain age were strongly correlated, including both in the same model caused collinearity issues and resulted in loss of significance (**Supplementary Table S9)**.

#### Model Performance Comparison

We observed that models for predicting severe mass effect including atrophy showed improved calibration compared with the baseline model **(Supplementary Figure 9).**

When stratified by admission era (pre-2015: n=331; post-2015: n=234), adding atrophy improved discrimination in both groups. In the pre-2015 cohort, AUC increased from 0.60 (baseline) to 0.67 (baseline + atrophy). In the post-2015 cohort, AUC increased from 0.62 to 0.70 respectively (**Supplementary Table S10**).

We observed that models including relative brain age had significantly improved goodness of fit compared to the baseline model (LRT $\chi^2$ (1) = 41; $p < 0.001$). They also had better balance between goodness of fit and model complexity as reflected by lower AIC and BIC values (AIC 703 vs. 741; BIC 733 vs. 767), modestly higher discrimination (AUC 0.67 vs. 0.60), and improved calibration/accuracy (Brier score 0.22 vs. 0.23; Mean Absolute Error 0.01 vs. 0.03) compared to the baseline model. There was no improvement in predicting mortality (**Supplementary Table S11, Supplementary Figures S10-S11**).

#### Other Exploratory Outcomes

Linear regression model comparison confirmed that including volumetric measures improved model fit for maximum midline shift within 10 days of stroke onset (LRT $\chi^2$ (1) = 238; p = <0.001), with adjusted $R^2$ increasing from 0.027 to 0.29 and root mean squared error decreasing from 4.01 to 3.96. It improved model fit for early midline shift within 24 hours of stroke onset LRT $\chi^2$ (1) = 84; p = <0.001), with adjusted $R^2$ increasing from 0.06 to 0.08 and root mean squared error decreasing from 2.44 to 2.42. (**Supplementary Table S12**).

**Supplementary Figures and Figure Legends**

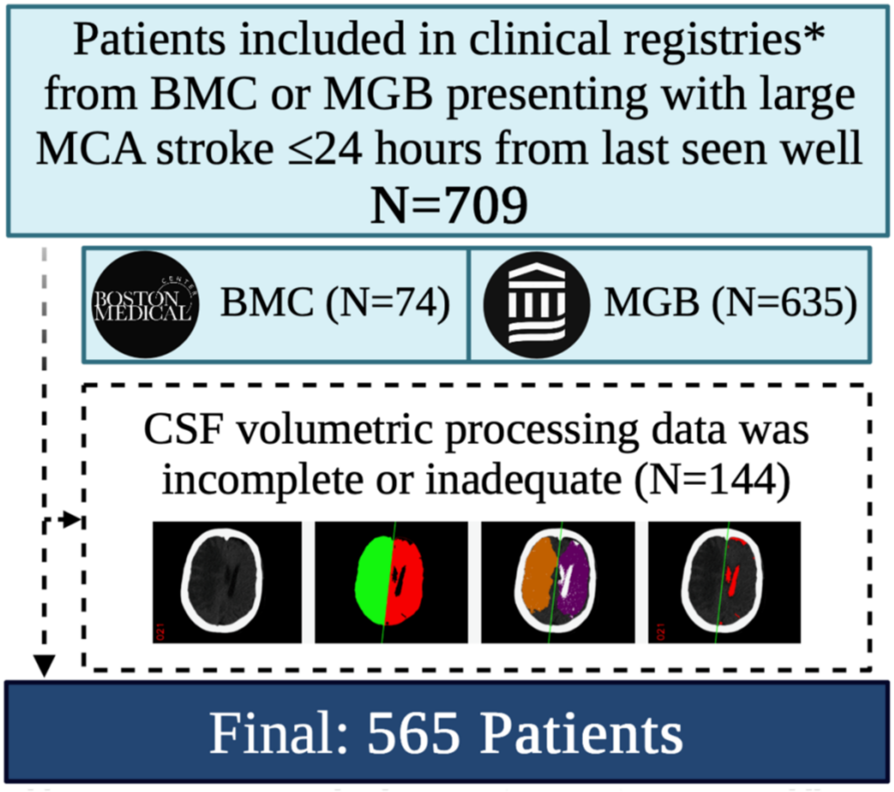

**Figure S1**. Patient eligibility. *Clinical registries included CNS and QUIP IRB registries. BMC=Boston Medical Center (2019-2024); CNS = Diagnosis and Management of Central Nervous System Lesions in the Neuro ICU: Identifying Differentiating Features and Predictors of Improved Outcomes, in order to Improve Quality of Care (IRB 2017P002564); CSF=cerebrospinal fluid; MCA=Middle Cerebral Artery; MGB=Mass General Brigham (2006-2018); QUIP = Quantitative Pupillometry in Patients with Critical Neurologic Injury (IRB H-37699).

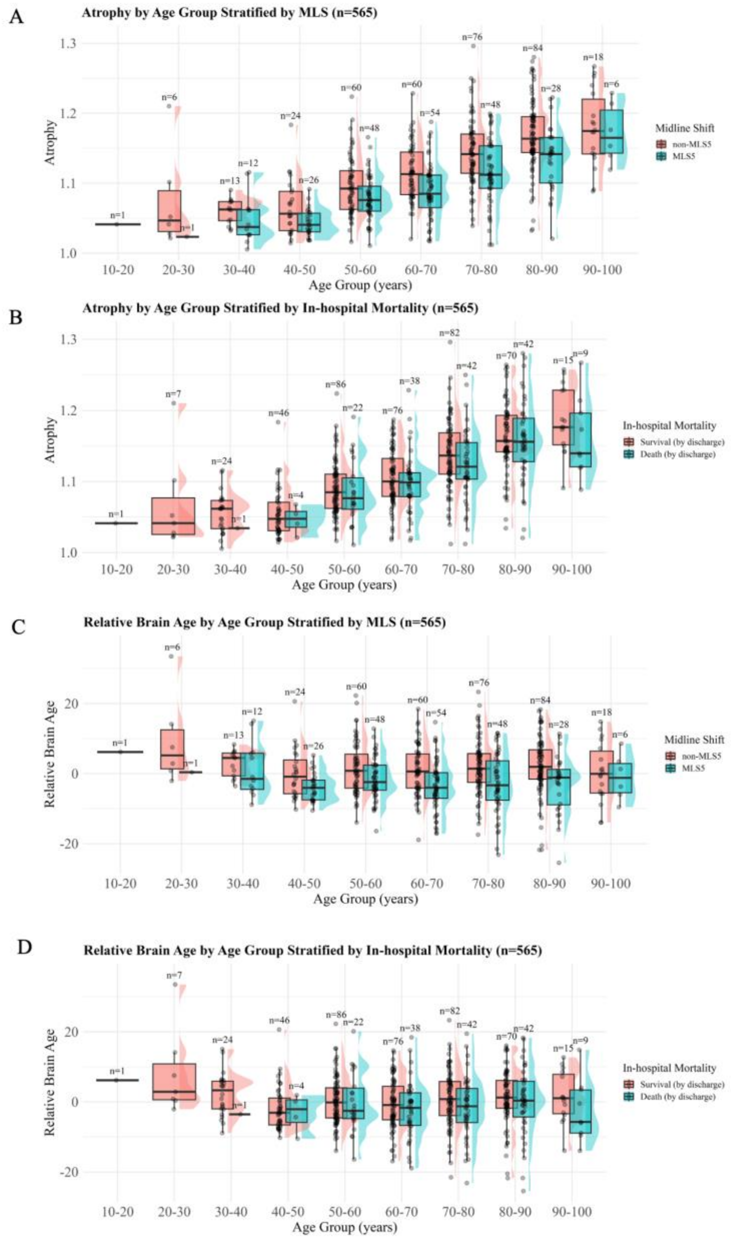

**Figure S2**. Distributions of atrophy and relative brain age across 10-year age groups, stratified by midline shift ≥5 mm and in-hospital mortality. (A) Atrophy increases with age, and the midline shift ≥5 mm group shows lower atrophy across most age bands. (B) Atrophy is similar by mortality across ages. (C) Relative brain age is independent of age, and the midline shift ≥5 mm group shows lower relative brain age (younger appearing brains) across most age bands. (D) Relative brain age shows no consistent difference by mortality. Boxes show medians and IQRs; violins, kernel densities; dots, individual patients. MLS=Midline Shift.

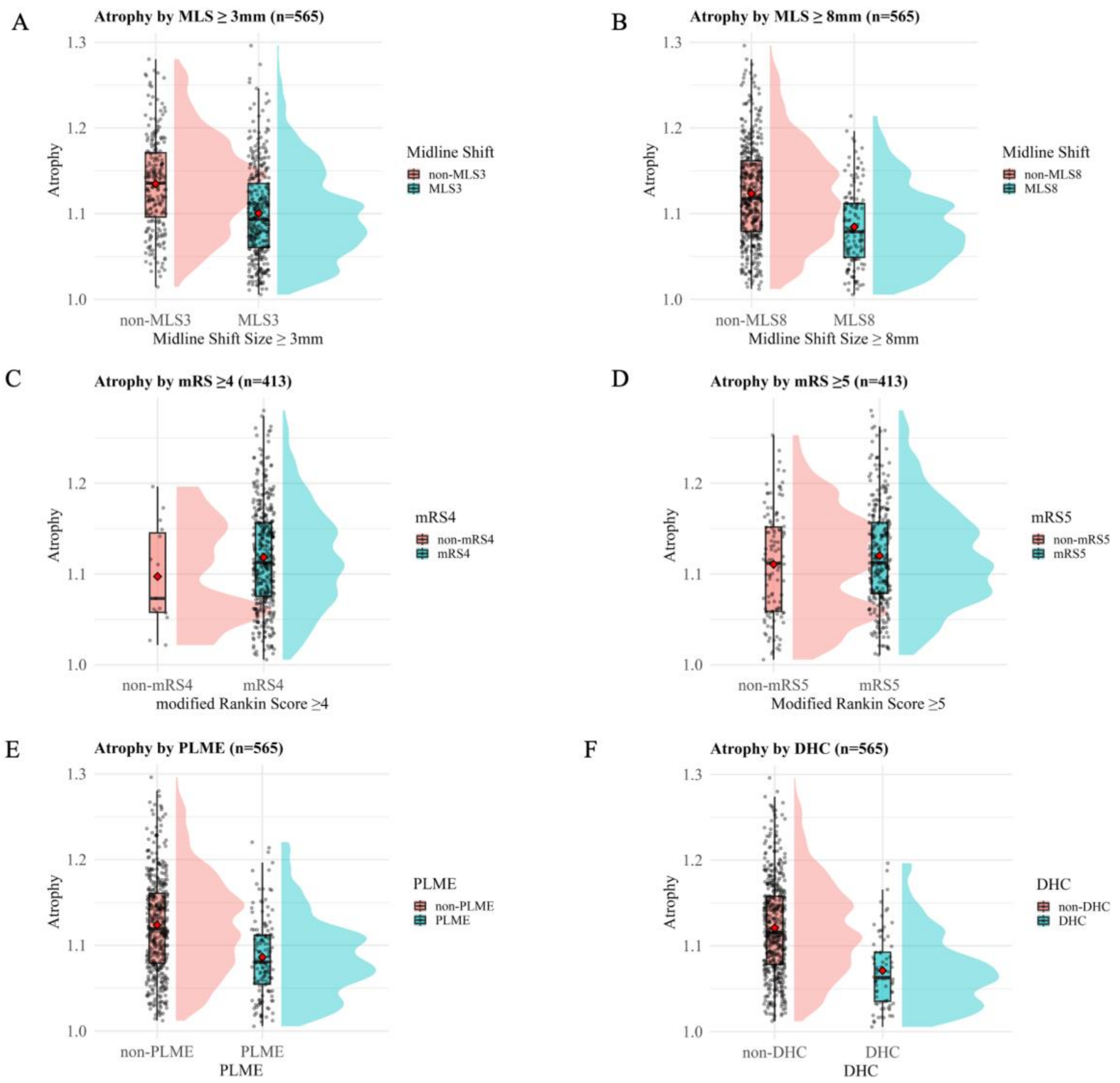

**Figure S3**. Comparisons of atrophy by other clinical phenotypes. The raincloud plots demonstrate decreased atrophy in patients with (A) midline shift ≥3mm and (B) midline shift ≥8mm, (E) potential lethal mass effect, and (F) decompressive hemicraniectomy. No apparent differences between patients with and without (C) modified Rankin Score ≥4 and (D) modified Rankin Score ≥5. MLS = midline shift; mRS = modified Rankin Score; PLME = potential lethal mass effect; DHC = decompressive hemicraniectomy.

A

B

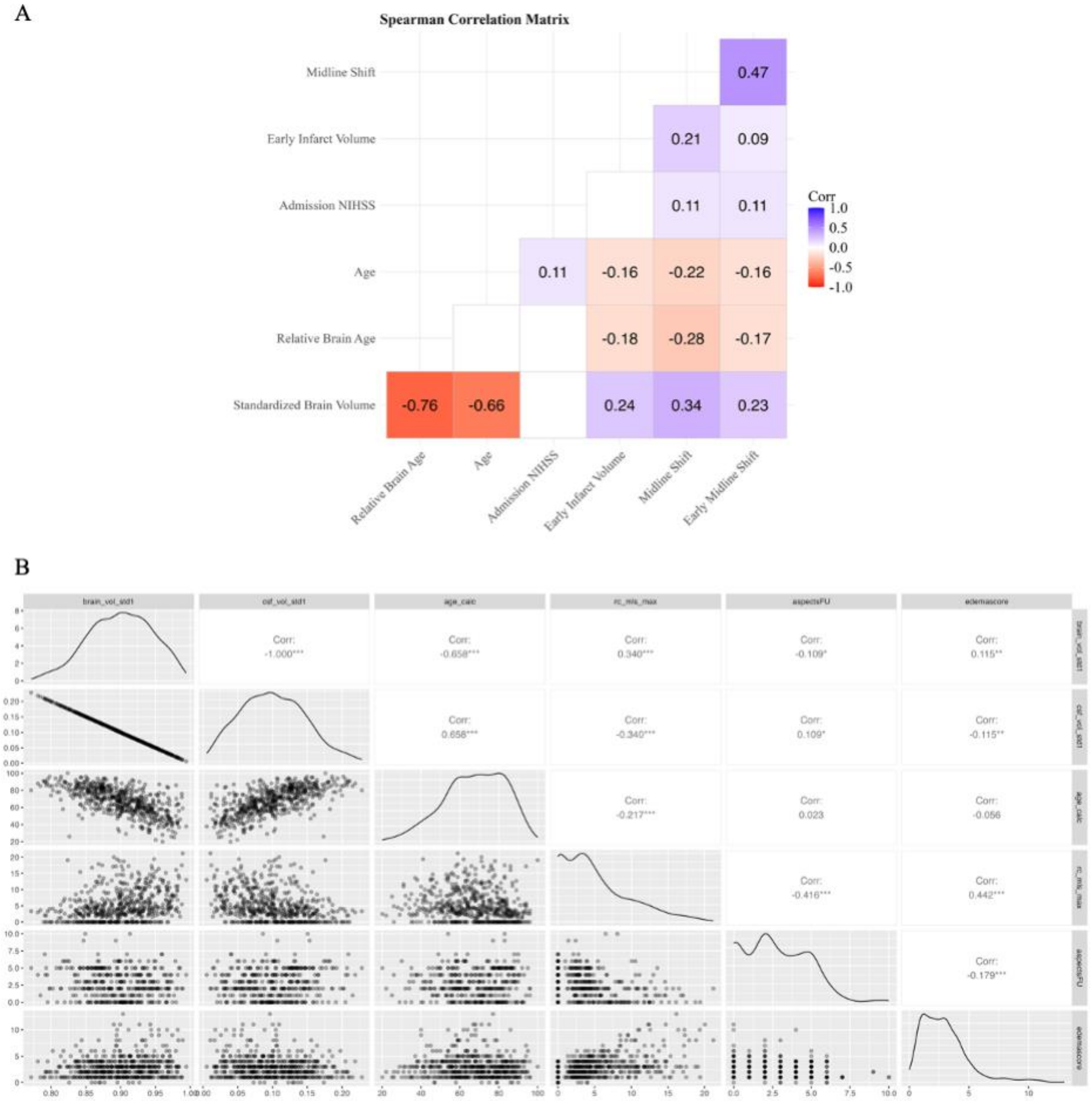

**Figure S4.** Correlations of atrophy and relative brain age with other continuous variables. (A) heatmap shows the pair-wise spearman correlations. Red boxes indicate negative correlations and purple boxes indicate positive correlations, with color intensity indicating correlation strength. (B) correlation matrix shows the scatter plots and correlation coefficients and their significance.

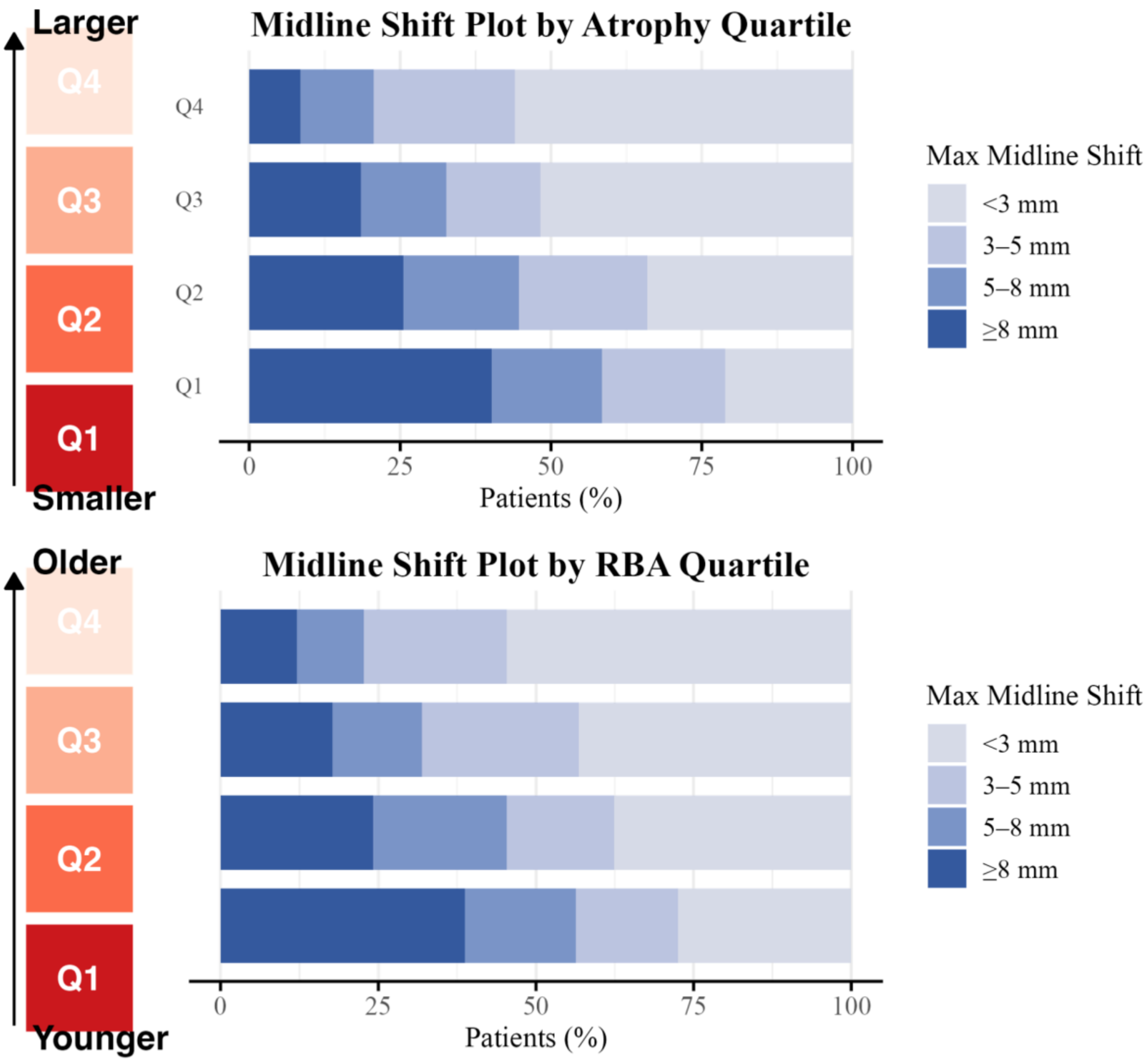

**Figure S5.** Distributions of maximum midline shift by quartiles of atrophy and relative brain age. There was a smaller proportion of severe mass effect (midline shift ≥5mm) in patients with higher quartiles of (A) atrophy and (B) relative brain age. Q1-Q4 quartiles of atrophy, relative brain age and maximum midline shift. Q1-Q2 represents patients with smaller atrophy, lower relative brain age (younger-appearing brains), whereas Q3-Q4 represents greater atrophy and older-looking brains. RBA = relative brain age.

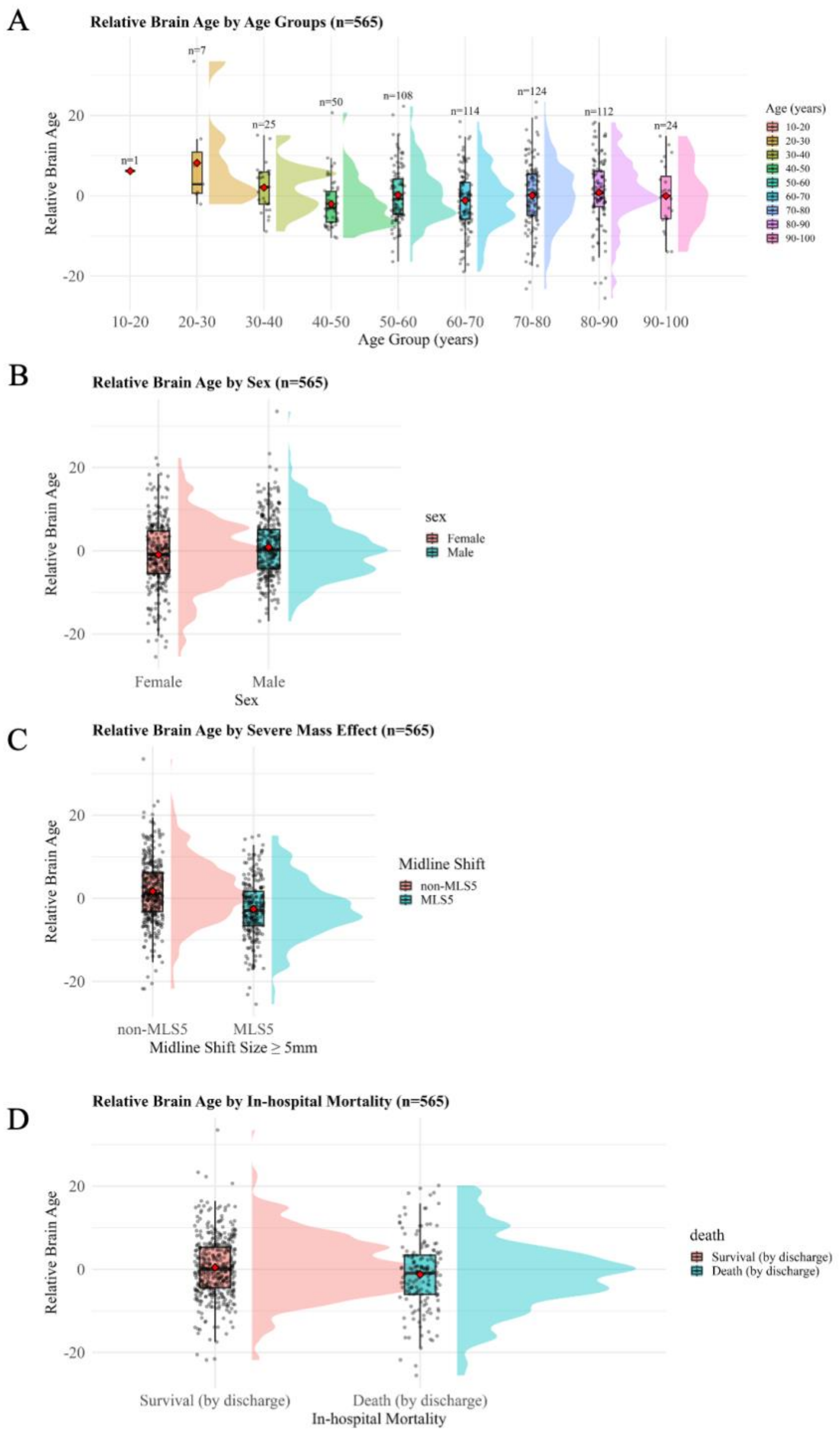

**Figure S6.** Relative brain age stratified by age, sex, midline shift, and in-hospital mortality. Patients with (A) younger age and those with (C) midline shift ≥5 mm tend to have lesser atrophy and younger relative brain age. No apparent differences in atrophy or relative brain age are observed across (B) sex or (D) in-hospital mortality.

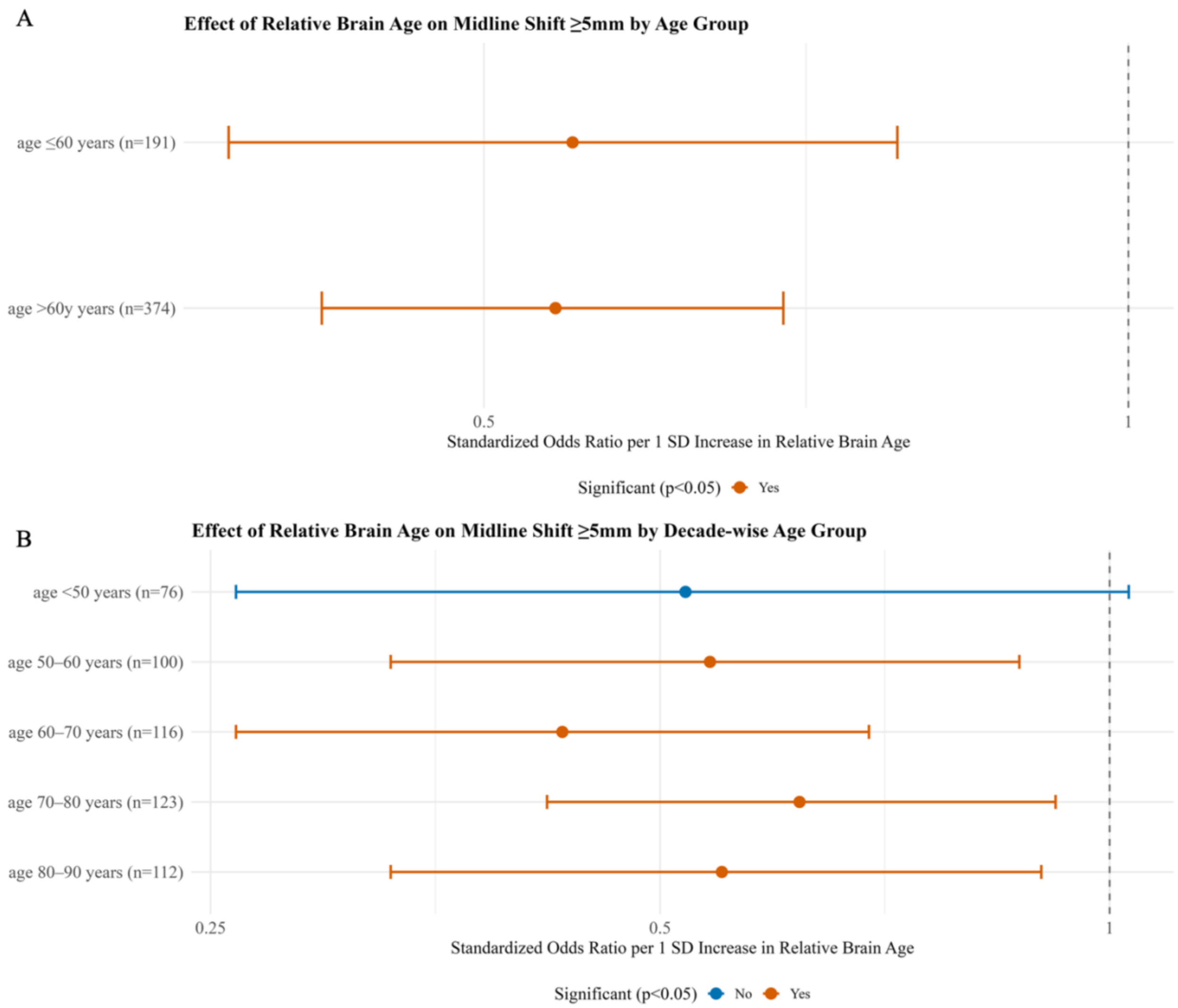

**Figure S7.** Effect size of relative brain age for severe mass effect, (A) dichotomized into two age groups and (B) stratified into 10-year age groups. The strongest protective effect of older relative brain age occurs in the 60–70-year group. Age groups <50 years (n=76) and ≥90 years (n=24) were excluded due to insufficient sample size for reliable estimation. The minimum sample size required to detect a significant effect ($p$<0.05) of a 1-SD change in relative brain age with 95% power is 112. Error bars represent 95% confidence intervals**.** SD = standard deviation.

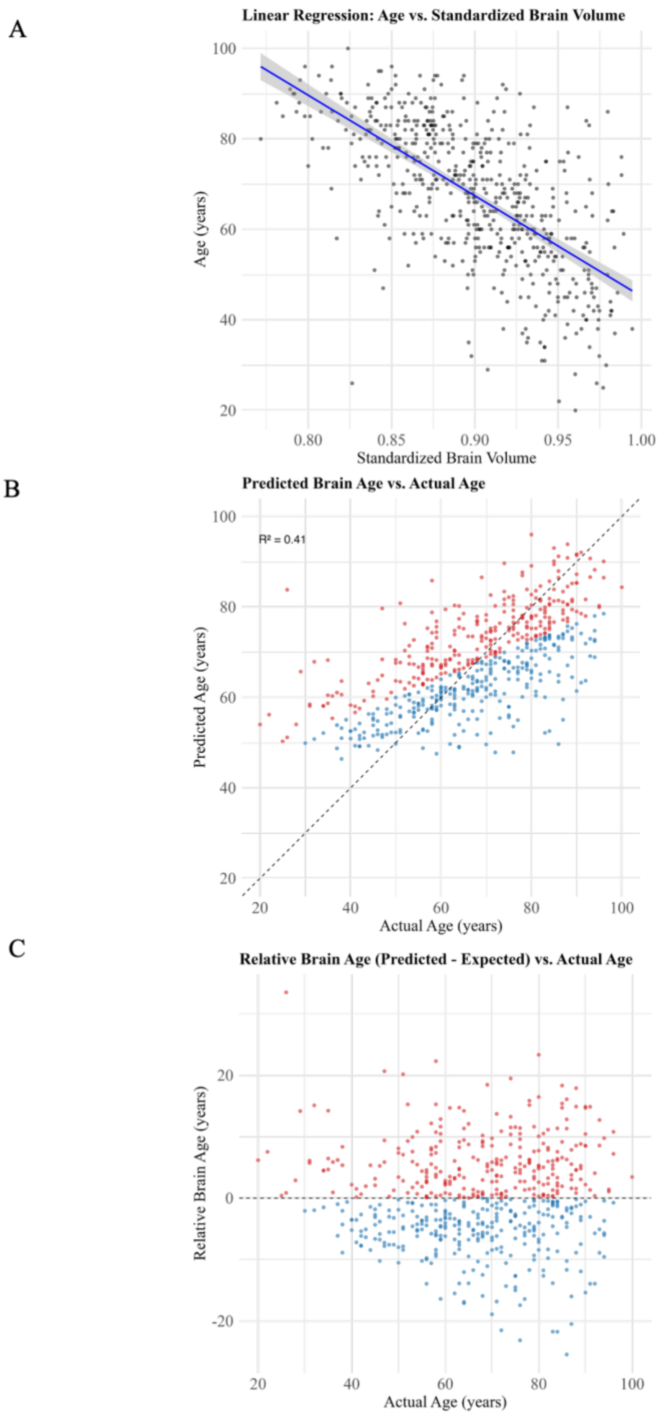

**Figure S8.** Brain age and relative brain age correlations. (A) Standardized brain volume is strongly negatively correlated with age. (B) Predicted brain age is strongly correlated with actual age. (C) Relative brain age is independent of actual age. Blue points indicate a predicted brain age younger than expected brain age, whereas red points indicate a predicted brain age older than expected brain age.

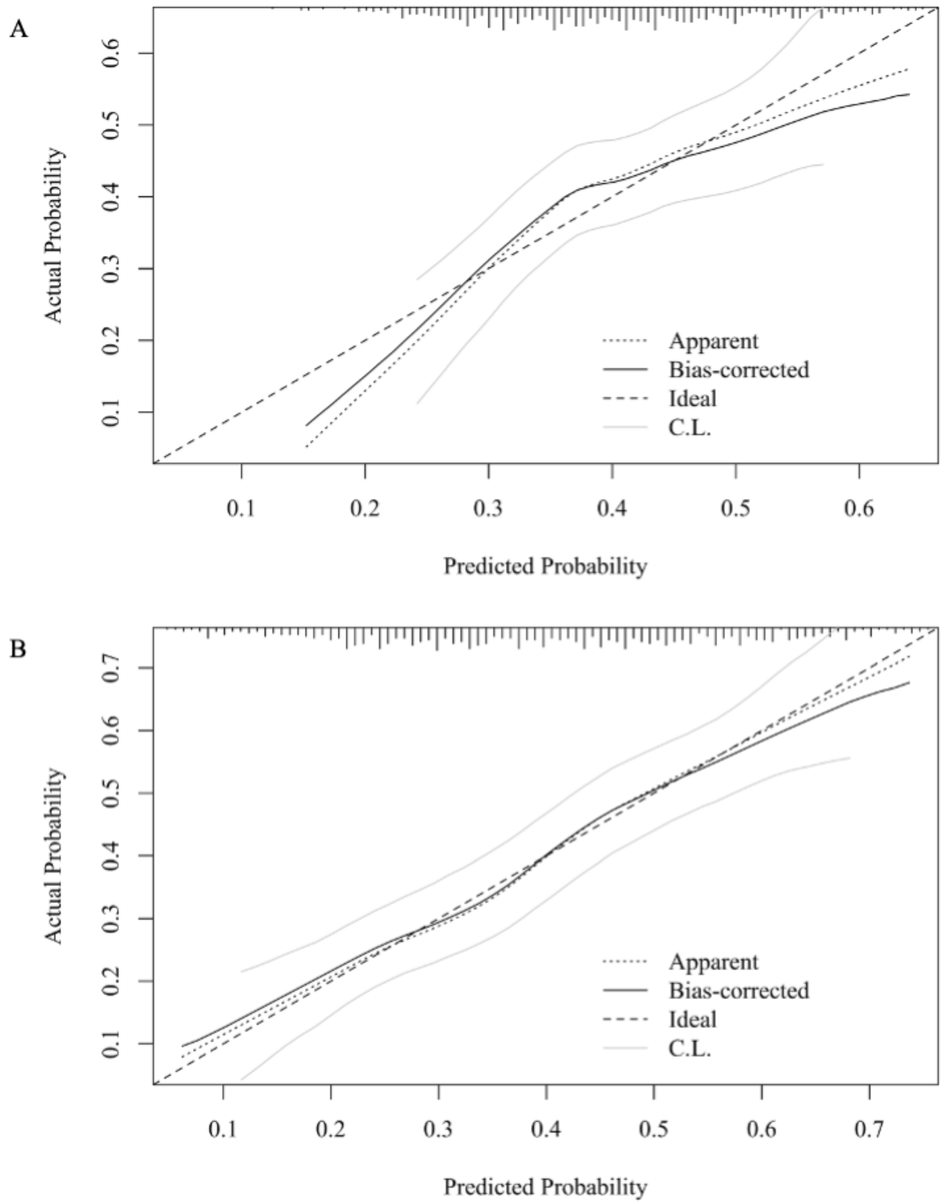

**Figure S9.** Model comparison of severe mass effect by calibration plots. Apparent (dotted lines) and bias-corrected (solid lines) calibration curves are plotted against the ideal line (dashed). Bias correction was performed using bootstrap resampling with 200 iterations. Models including atrophy show improved calibration compared with (A) the baseline model, as indicated by closer alignment of the bias-corrected curve with the ideal line.

A

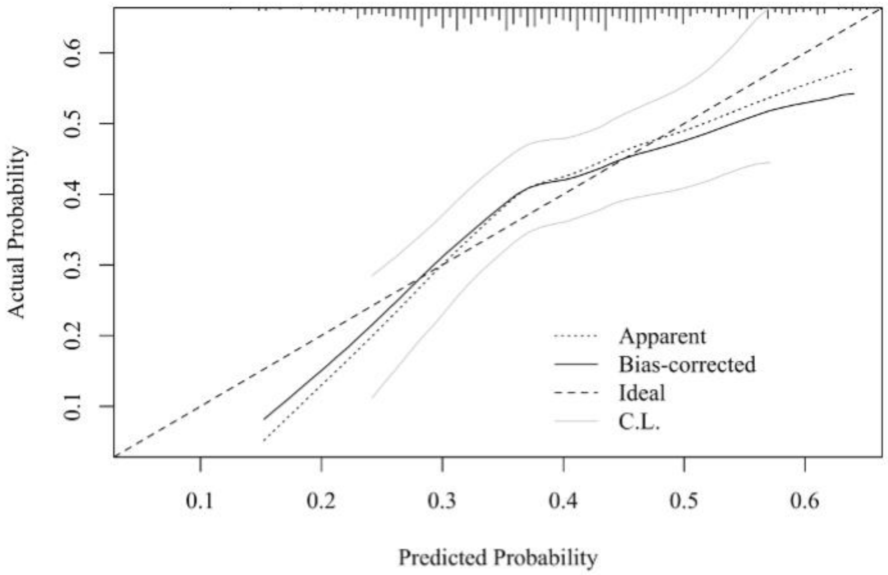

B

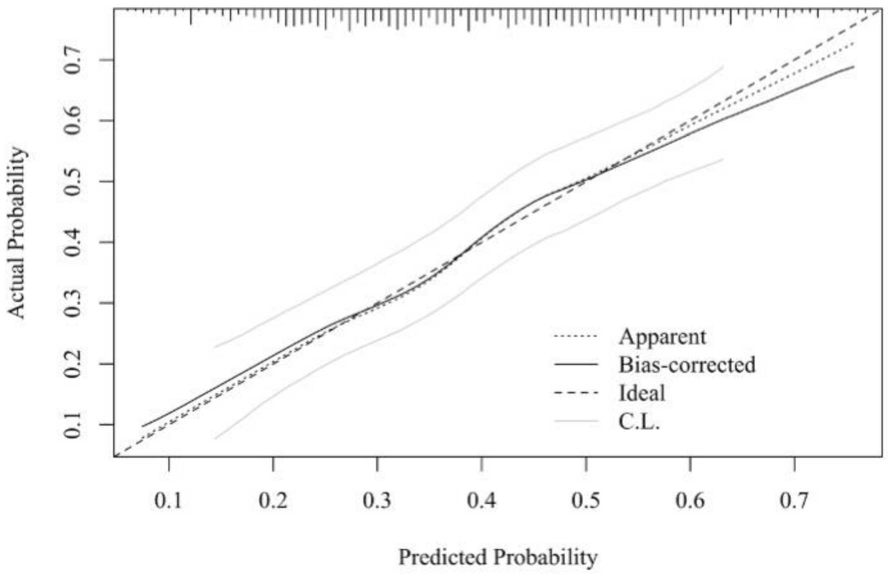

**Figure S10.** Model comparison of severe mass effect by calibration plots. Apparent (dotted lines) and bias-corrected (solid lines) calibration curves are plotted against the ideal line (dashed). Bias correction was performed using bootstrap resampling with 200 iterations. Models including either (B) relative brain age show improved calibration compared with (A) the baseline model, as indicated by closer alignment of the bias-corrected curve with the ideal line.

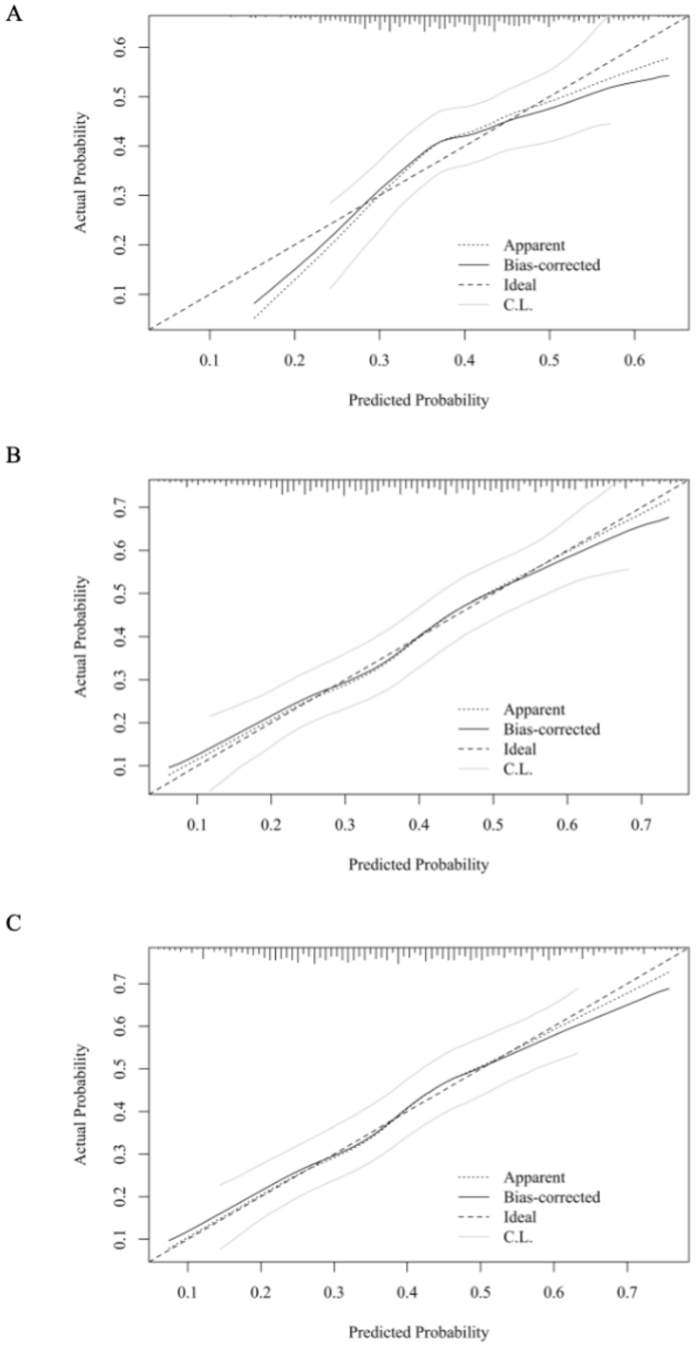

**Figure S11.** Model comparison of in-hospital mortality by calibration plots. Apparent (dotted lines) and bias-corrected (solid lines) calibration curves are plotted against the ideal line (dashed). Bias correction was performed using bootstrap resampling with 200 iterations. Models including either (B) atrophy or (C) relative brain age show no improved calibration compared with (A) the baseline model, as indicated by same alignment of the bias-corrected curve with the ideal line.

#### Supplementary Tables

**Table S1.** Missing Data Summary

| Variable | N Total | N Observed | N Missing | Percentage Missing (%) |
|---|---|---|---|---|
| **Age (years)** | 565 | 565 | 0 | 0.0 |
| **Admission NIHSS** | 565 | 526 | 39 | 6.9 |
| **Hyperglycemia**[a] | 565 | 563 | 2 | 0.4 |
| **Intervention**[b] | 565 | 565 | 0 | 0.0 |
| **Infarct Volume**[c] | 565 | 563 | 2 | 0.4 |
| **Infarct Volume before MLS5**[d] | 565 | 510 | 55 | 9.7 |
| **Infarct Volume before MLS3**[d] | 565 | 450 | 115 | 20.4 |
| **Infarct Volume before MLS8**[d] | 565 | 543 | 22 | 3.9 |
| **Infarct Volume before PLME**[d] | 565 | 562 | 3 | 0.5 |
| **Infarct Volume before DHC**[d] | 565 | 562 | 3 | 0.5 |

[a.] Binary indicator of whether glucose ≥150 mg/dL. [b.] Includes intravenous thrombolysis or mechanical thrombectomy. [c.] Defined as maximum infarct volume measured on head CT within 10 days of last seen well. Discharge outcomes and continuous outcomes were adjusted by this covariate. [d.] Defined as maximum infarct volume measured on head CT before event occurrence. DHC=Decompressive Hemicraniectomy; MLS5=Midline Shift ≥5 mm; MLS3=Midline Shift ≥3 mm; MLS8=Midline Shift ≥8 mm; NIHSS=National Institutes of Health Stroke Scale; PLME=Potentially Lethal Malignant Edema.

**Table S2.** Effect Size of Atrophy for Other Binary Clinical Outcomes

| | Standardized Odds Ratio[a] | | | |
|---|---|---|---|---|
| | | 95% Confidence Intervals | | |
| **Outcomes** | **Odds Ratio** | **Lower** | **Upper** | **p Value** |
| **Secondary Outcome** | | | | |
| In-hospital Mortality | 0.87 | 0.67 | 1.14 | 0.320 |
| **Exploratory Outcomes** | | | | |
| Midline Shift ≥3 mm | 0.55 | 0.43 | 0.70 | <0.001 |
| Midline Shift ≥8 mm | 0.40 | 0.29 | 0.55 | <0.001 |
| Potentially Lethal Malignant Edema | 0.57 | 0.41 | 0.78 | <0.001 |
| Decompressive Hemicraniectomy | 0.57 | 0.37 | 0.88 | 0.010 |
| mRS ≥4 at Discharge | 1.68 | 0.73 | 3.86 | 0.221 |
| mRS ≥5 at Discharge | 0.92 | 0.67 | 1.26 | 0.593 |
| **Subgroup Analysis** | | | | |
| Midline Shift ≥5 mm in Patients without PH2[b] (n = 530) | 0.41 | 0.31 | 0.55 | <0.001 |
| Midline Shift ≥5 mm in MGB Patients only (n = 491) | 0.48 | 0.36 | 0.64 | <0.001 |
| Midline Shift ≥5 mm in patients admitted before 2015 (n = 331) | 0.52 | 0.36 | 0.74 | <0.001 |
| Midline Shift ≥5 mm in patients admitted after 2015 (n = 234) | 0.40 | 0.26 | 0.60 | <0.001 |

[a]Standardized odds ratios quantified the effects of a 1 standard deviation increase in each continuous predictor. [b]PH2 defined as hematoma >30% of infarct zone, associated with substantial mass effect. MGB = Mass General Brigham Hospital; mRS = modified Rankin Scale; NIHSS = National Institutes of Health Stroke Scale; PH2 = Parenchymal Hemorrhage subtype 2.

**Table S3.** Linear Regression of Maximum Midline Shift

| Variables | Coefficient | 95% Confidence Intervals | | p Value |
| --- | --- | --- | --- | --- |
| | | Lower | Upper | |
| **Baseline Model** | | | | |
| Age | -0.031 | -0.052 | -0.009 | 0.006 |
| NIHSS | 0.030 | -0.029 | 0.089 | 0.314 |
| Hyperglycemia | 0.863 | 0.145 | 1.582 | 0.019 |
| Intervention | 0.391 | -0.282 | 1.064 | 0.255 |
| Early Infarct Volume | 0.025 | 0.022 | 0.029 | <0.001 |
| **Baseline Model with Atrophy** | | | | |
| Age | 0.003 | -0.024 | 0.031 | 0.819 |
| NIHSS | 0.029 | -0.029 | 0.087 | 0.324 |
| Hyperglycemia | 0.791 | 0.081 | 1.502 | 0.029 |
| Intervention | 0.261 | -0.407 | 0.930 | 0.443 |
| Early Infarct Volume | 0.023 | 0.020 | 0.027 | <0.001 |
| Atrophy | -15.115 | -22.904 | -7.326 | <0.001 |
| **Baseline Model with Relative Brain Age** | | | | |
| Age | -0.032 | -0.053 | -0.011 | 0.004 |
| NIHSS | 0.029 | -0.029 | 0.087 | 0.322 |
| Hyperglycemia | 0.793 | 0.084 | 1.503 | 0.029 |
| Intervention | 0.251 | -0.417 | 0.919 | 0.461 |
| Early Infarct Volume | 0.023 | 0.019 | 0.027 | <0.001 |
| Relative Brain Age | -0.088 | -0.133 | -0.044 | <0.001 |

[a]Maximum midline shift was evaluated within 10 days of stroke onset. NIHSS = National Institutes of Health Stroke Scale.

**Table S4.** Linear Regression of Early Maximum Midline Shift

| Variables | Coefficient | 95% Confidence Intervals | | p Value |
|---|---|---|---|---|
| | | Lower | Upper | |
| **Baseline Model** | | | | |
| Age | -0.015 | -0.029 | -0.002 | 0.023 |
| NIHSS | 0.032 | -0.004 | 0.068 | 0.080 |
| Hyperglycemia | 0.213 | -0.225 | 0.651 | 0.339 |
| Intervention | 0.428 | 0.017 | 0.839 | 0.041 |
| Early Infarct Volume | 0.006 | 0.003 | 0.008 | <0.001 |
| **Baseline Model with Atrophy** | | | | |
| Age | 0.004 | -0.013 | 0.020 | 0.673 |
| NIHSS | 0.031 | -0.004 | 0.067 | 0.085 |
| Hyperglycemia | 0.172 | -0.262 | 0.606 | 0.436 |
| Intervention | 0.354 | -0.055 | 0.763 | 0.089 |
| Early Infarct Volume | 0.004 | 0.002 | 0.007 | <0.001 |
| Atrophy | -8.498 | -13.254 | -3.742 | <0.001 |
| **Baseline Model with Relative Brain Age** | | | | |
| Age | -0.016 | -0.029 | -0.003 | 0.015 |
| NIHSS | 0.031 | -0.004 | 0.067 | 0.084 |
| Hyperglycemia | 0.171 | -0.262 | 0.604 | 0.437 |
| Intervention | 0.344 | -0.064 | 0.752 | 0.099 |
| Early Infarct Volume | 0.004 | 0.002 | 0.007 | <0.001 |
| Relative Brain Age | -0.052 | -0.080 | -0.025 | <0.001 |

* Early maximum midline shift was evaluated within 24 hours of stroke onset. NIHSS = National Institutes of Health Stroke Scale

**Table S5.** Effect Size of Atrophy Stratified by Age

| Outcomes | Standardized Odds Ratio[a] | | | p Value |
|---|---|---|---|---|
| | | 95% Confidence Intervals | | |
| | Odds Ratio | Lower | Upper | |
| **Subgroup Analysis with Binary Age** | | | | |
| Midline Shift ≥5 mm in patients aged ≤60 years (n = 191) | 0.52 | 0.36 | 0.77 | <0.001 |
| Midline Shift ≥5 mm in patients aged >60 years (n = 374) | 0.49 | 0.36 | 0.65 | <0.001 |
| **Subgroup Analysis with Decade-wise Age** | | | | |
| Midline Shift ≥5 mm in patients aged <50 years (n = 76) | 0.55 | 0.29 | 1.05 | 0.069 |
| Midline Shift ≥5 mm in patients aged 50-60 years (n = 100) | 0.53 | 0.32 | 0.87 | 0.012 |
| Midline Shift ≥5 mm in patients aged 60-70 years (n = 116) | 0.43 | 0.26 | 0.70 | <0.001 |
| Midline Shift ≥5 mm in patients aged 70-80 years (n = 123) | 0.62 | 0.41 | 0.93 | 0.020 |
| Midline Shift ≥5 mm in patients aged 80-90 years (n = 112) | 0.52 | 0.30 | 0.89 | 0.018 |
| Midline Shift ≥5 mm in patients aged >90 years (n = 24) | - | - | - | - |

[a]Standardized odds ratios quantified the effects of a 1 standard deviation increase in each continuous predictor. Age group ≥90 years (n=24) has no reliable estimation due to insufficient sample size. The minimum sample size required to detect a significant effect ($p$<0.05) of a 1-SD change in atrophy with 80% power is 50. SD = standard deviation.

**Table S6.** Logistic Regression of Relative Brain Age and Severe Mass Effect

| | Standardized Odds Ratio* | | | |
|---|---|---|---|---|
| | | 95% Confidence Intervals | | |
| **Variables** | **Odds Ratio** | **Lower** | **Upper** | **p Value** |
| **Baseline Model** | | | | |
| Age | 0.69 | 0.58 | 0.83 | <0.001 |
| NIHSS | 1.29 | 1.08 | 1.55 | 0.006 |
| Hyperglycemia | 1.51 | 1.04 | 2.19 | 0.031 |
| Intervention | 1.03 | 0.72 | 1.46 | 0.889 |
| Early Infarct Volume | 1.08 | 0.90 | 1.28 | 0.404 |
| **Baseline Model with Relative Brain Age** | | | | |
| Age | 0.67 | 0.56 | 0.81 | <0.001 |
| NIHSS | 1.29 | 1.07 | 1.55 | 0.009 |
| Hyperglycemia | 1.45 | 0.98 | 2.13 | 0.062 |
| Intervention | 0.90 | 0.62 | 1.30 | 0.564 |
| Early Infarct Volume | 1.00 | 0.83 | 1.21 | 0.975 |
| Relative Brain Age | 0.54 | 0.45 | 0.66 | <0.001 |

* Standardized odds ratios quantified the effects of a 1 standard deviation increase in each continuous predictor. Standard deviation for each continuous predictors: age, 15.7 years; NIHSS, 5.7; Early Infarct Volume, 71.1 mL; Relative Brain Age, 7.7 years.
NIHSS = National Institutes of Health Stroke Scale

**Table S7.** Effect Size of Relative Brain Age for Other Binary Clinical Outcomes

| | Standardized Odds Ratio[a] | | | |
|---|---|---|---|---|
| | | 95% Confidence Intervals | | |
| **Outcomes** | **Odds Ratio** | **Lower** | **Upper** | **p Value** |
| **Secondary Outcome** | | | | |
| In-hospital Mortality | 0.90 | 0.73 | 1.10 | 0.298 |
| **Exploratory Outcomes** | | | | |
| Midline Shift ≥3 mm | 0.62 | 0.51 | 0.75 | <0.001 |
| Midline Shift ≥8 mm | 0.51 | 0.41 | 0.65 | <0.001 |
| Potentially Lethal Malignant Edema | 0.66 | 0.52 | 0.83 | <0.001 |
| Decompressive Hemicraniectomy | 0.66 | 0.49 | 0.90 | 0.009 |
| mRS ≥4 at Discharge | 1.50 | 0.80 | 2.82 | 0.208 |
| mRS ≥5 at Discharge | 0.94 | 0.74 | 1.20 | 0.633 |
| **Subgroup Analysis** | | | | |
| Midline Shift ≥5 mm in Patients without PH2[b] (n = 530) | 0.52 | 0.42 | 0.65 | <0.001 |
| Midline Shift ≥5 mm in MGB Patients only (n = 491) | 0.59 | 0.48 | 0.73 | <0.001 |
| Midline Shift ≥5 mm in patients admitted before 2015 (n = 331) | 0.62 | 0.48 | 0.81 | <0.001 |
| Midline Shift ≥5 mm in patients admitted after 2015 (n = 234) | 0.48 | 0.35 | 0.67 | <0.001 |

[a]Standardized odds ratios quantified the effects of a 1 standard deviation increase in each continuous predictor. [b]PH2 defined as hematoma >30% of infarct zone, associated with substantial mass effect. MGB = Mass General Brigham Hospital; mRS = modified Rankin Scale; NIHSS = National Institutes of Health Stroke Scale; PH2 = Parenchymal Hemorrhage subtype 2.

**Table S8.** Effect Size of Relative Brain Age Stratified by Age

| | Standardized Odds Ratio[a] | | | |
|---|---|---|---|---|
| | | 95% Confidence Intervals | | |
| **Outcomes** | **Odds Ratio** | **Lower** | **Upper** | **p Value** |
| **Subgroup Analysis with Binary Split** | | | | |
| Midline Shift ≥5 mm in patients aged ≤60 years (n = 191) | 0.55 | 0.38 | 0.78 | <0.001 |
| Midline Shift ≥5 mm in patients aged >60 years (n = 374) | 0.54 | 0.42 | 0.69 | <0.001 |
| **Subgroup Analysis with Decade-wise Age** | | | | |
| Midline Shift ≥5 mm in patients aged <50 years (n = 76) | 0.52 | 0.26 | 1.03 | 0.062 |
| Midline Shift ≥5 mm in patients aged 50-60 years (n = 100) | 0.54 | 0.33 | 0.87 | 0.011 |
| Midline Shift ≥5 mm in patients aged 60-70 years (n = 116) | 0.43 | 0.26 | 0.69 | <0.001 |
| Midline Shift ≥5 mm in patients aged 70-80 years (n = 123) | 0.62 | 0.42 | 0.92 | 0.018 |
| Midline Shift ≥5 mm in patients aged 80-90 years (n = 112) | 0.55 | 0.33 | 0.90 | 0.019 |
| Midline Shift ≥5 mm in patients aged>90 years (n = 24) | - | - | - | - |

[a] Standardized odds ratios quantified the effects of a 1 standard deviation increase in each continuous predictor. The protective effect of older relative brain age is significant only in patients older than 50 years. The strongest protective effect occurs in the 60–70-year group. Age groups <50 years (n=76) and ≥90 years (n=24) have no reliable estimation due to insufficient sample size. The minimum sample size required to detect a significant effect ($p$<0.05) of a 1-SD change in relative brain age with 80% power is 9**8**. SD = standard deviation.

**Table S9.** Logistic Regression of Severe Mass Effect including Combinations of Chronological age, Relative Brain Age and Atrophy

| | Standardized Odds Ratio[a] | | | |
|---|---|---|---|---|
| | | 95% Confidence Intervals | | |
| **Variables** | **Odds Ratio** | **Lower** | **Upper** | **p Value** |
| **A) Base Model with Relative Brain Age and Atrophy** | | | | |
| Age | 1.28 | 0.13 | 12.17 | 0.829 |
| NIHSS | 1.29 | 1.07 | 1.55 | 0.009 |
| Hyperglycemia | 1.44 | 0.98 | 2.12 | 0.064 |
| Intervention | 0.90 | 0.62 | 1.30 | 0.574 |
| Early Infarct Volume | 1.00 | 0.83 | 1.21 | 0.990 |
| Relative Brain Age | 1.18 | 0.08 | 17.58 | 0.902 |
| Atrophy | 0.36 | 0.01 | 12.74 | 0.572 |
| **B) Base Model (without age) with Relative Brain Age and Atrophy** | | | | |
| NIHSS | 1.29 | 1.07 | 1.55 | 0.009 |
| Hyperglycemia | 1.44 | 0.98 | 2.13 | 0.063 |
| Intervention | 0.90 | 0.62 | 1.30 | 0.570 |
| Early Infarct Volume | 1.00 | 0.83 | 1.21 | 0.986 |
| Relative Brain Age | 0.88 | 0.66 | 1.17 | 0.388 |
| Atrophy | 0.53 | 0.39 | 0.71 | <0.001 |

Logistic regression models including age, relative brain age, and atrophy are shown for completeness. As both relative brain age and atrophy are derived from standardized brain volume, collinearity caused unstable estimates and loss of significance. These findings support our focus on the *age + relative brain age* and *age + atrophy* models in the main analysis. [a] Standardized odds ratios quantified the effects of a 1-standard deviation increase in each continuous predictor. NIHSS = National Institutes of Health Stroke Scale.

**Table S10**. Comparison of Logistic Regression Models for Severe Mass Effect with and without Atrophy Stratified by Admission Year

| Outcome | Admission Year | Model | Model Fit | | | | Discrimination | | Calibration Accuracy Statistics |
|---|---|---|---|---|---|---|---|---|---|
| | | | Likelihood Ratio Test | | | | | | |
| | | | $\chi^2$ statistic | p Value | AIC | BIC | AUC | Brier Score | Mean Absolute Error |
| Severe Mass Effect | Pre-2015 (n = 331) | Baseline | | | 421.97 | 444.78 | 0.60 | 0.22 | 0.02 |
| | | Baseline + Atrophy | 14.26 | <0.001 | 409.71 | 436.33 | 0.67 | 0.21 | 0.02 |
| | Post-2015 (n = 234) | Baseline | | | 316.03 | 336.76 | 0.62 | 0.24 | 0.04 |
| | | Baseline + Atrophy | 22.27 | <0.001 | 295.76 | 319.95 | 0.70 | 0.23 | 0.03 |

* All models were adjusted by age, admission NIHSS, hyperglycemia, whether the patients have intravenous thrombolysis or mechanical thrombectomy, max infarct volume before event.

**Table S11**. Comparison of Logistic Regression Models with and without Relative Brain Age

| Outcomes | Model | Model Fit | | | | Discrimination | | Calibration Accuracy Statistics |
|---|---|---|---|---|---|---|---|---|
| | | Likelihood Ratio Test | | | | | | |
| | | $\chi^2$ statistic | p Value | AIC | BIC | AUC | Brier Score | Mean Absolute Error |
| Severe Mass Effect | Baseline | | | 741.37 | 767.39 | 0.60 | 0.23 | 0.03 |
| | Baseline + Relative Brain Age | 40.65 | <0.001 | 702.72 | 733.08 | 0.67 | 0.22 | 0.01 |
| Mortality by Discharge | Baseline | | | 606.58 | 632.60 | 0.71 | 0.18 | 0.01 |
| | Baseline + Relative Brain Age | 1.09 | 0.297 | 607.49 | 637.85 | 0.71 | 0.18 | 0.02 |

* All models were adjusted by age, admission NIHSS, hyperglycemia, whether the patients have intravenous thrombolysis or mechanical thrombectomy, max infarct volume before event.

**Table S12**. Comparison of Linear Regression Models with and without either Atrophy or Relative Brain Age

| Outcomes | Model | Model Fit | | | | | Calibration Accuracy Statistics |
|---|---|---|---|---|---|---|---|
| | | Likelihood Ratio Test | | | | | |
| | | $\chi^2$ statistic | p Value | AIC | BIC | Adjusted R-squared | Root Mean Squared Error |
| Max Midline Shift[a] | Baseline | | | 3177.68 | 3208.04 | 0.27 | 4.01 |
| | Baseline + Atrophy | 226.90 | <0.001 | 3165.16 | 3199.85 | 0.29 | 3.96 |
| | Baseline + Relative Brain Age | 238.22 | <0.001 | 3164.42 | 3199.12 | 0.29 | 3.96 |
| Early Max Midline Shift[b] | Baseline | | | 2608.00 | 2638.34 | 0.06 | 2.44 |
| | Baseline + Atrophy | 71.62 | <0.001 | 2597.66 | 2632.33 | 0.08 | 2.42 |
| | Baseline + Relative Brain Age | 83.803 | <0.001 | 2595.54 | 2630.20 | 0.08 | 2.42 |

All models were adjusted by age, admission NIHSS, Glucose ≥150, whether the patients have intravenous thrombolysis or mechanical thrombectomy, max infarct volume before event. [a] Maximum midline shift was evaluated within 10 days of stroke onset. [b]Early maximum midline shift was evaluated within 24 hours of stroke onset.
